# Genetic Variant Burden Rises with BMI: Insights from Rare and Low-Frequency Variants in Obesity

**DOI:** 10.64898/2026.10.08.26364979

**Authors:** Sumi Elsa John, Arshad Channanath, Md Zubbair Malik, Ashraf Al Madhoun, Fahd Al-Mulla, Thangavel Alphonse Thanaraj

## Abstract

Adult obesity, including morbid obesity, has reached epidemic proportions worldwide. Yet, the current genetic models based on BMI-associated common variants identified through genome-wide association studies (GWAS) explain only a modest fraction of BMI variability. Contribution of rare and low-frequency variants to polygenic obesity remains incompletely defined. Leveraging whole-exome sequences of 1138 adults spanning normal-weight, overweight, obese and morbidly obese (MO) categories from Kuwait and Qatar, we catalogued cohort-specific rare and low-frequency deleterious variants differentiating MO individuals from normal-weight individuals. Cross-cohort comparisons identified 147 risk variants seen in ≥2 MO individuals from one cohort and in ≥1 MO individual from the other; among these, 27 variants occurred in ≥2 MO individuals from each cohort. The risk genes were enriched in obesity-related pathways and tissues, with most harbouring GWAS variants associated primarily with obesity traits followed by metabolic and brain-related traits. Duodenal tissue expression data exhibited significant dysregulation of key risk genes in MO individuals. Variants from the 147-list and 27-list occurred in 87% and 49% of MO individuals, respectively; MO-variant burden correlated positively with BMI (r=0.30 and r=0.31, respectively) across the overweight-obese-MO categories. Combinations of a finite set of low-frequency coding variants substantially shape the continuum of polygenic obesity.

## Introduction

Obesity, characterized by excessive fat accumulation, has become a major global health epidemic. The Global Burden of Diseases, Injuries, and Risk Factors (GBD) Study considered rising overweight and obesity rates worldwide from 1990 to 2021, and projected that over half of all adults—about 3.8 billion people—will be affected by 2050^1^. A similar upward trend has been documented among children and adolescents over the past decades^2^. Obesity contributes to many medical comorbidities, such as type 2 diabetes (T2D), metabolic syndrome, sleep disorders and depression. The increasing prevalence of obesity significantly strains public health systems across the globe resulting in substantial healthcare costs, resource allocation, and long-term economic challenges for governmental healthcare providers. The economic impact of obesity affects society at large and underscores the urgent need for effective public health interventions^3–5^.

Morbid obesity represents a rapidly expanding subset of the obese population; for example, in the United States of America, the prevalence of severe obesity increased from 7.7% to 9,7% between 2013-2014 and 2021-2023^6^. In Kuwait, the mean onset age for T2D among morbidly obese individuals is reduced by 8.2 years (95% CI 5.2–11.2; *p* < 0.001) compared to normal-weight individuals^7^. While overweight and obese individuals may, in principle, achieve a return to normal-weight through dietary and physical activity interventions, morbidly obese individuals often require surgical and life-long pharmaceutical treatments.

Although environmental factors contribute to obesity prevalence, substantial evidence indicates that genetic predisposition plays a significant role. Identical twins, who share the same genetic makeup, exhibit more similar body weights than dizygotic twins. Obesity resulting from genetic causes are of two primary forms: monogenic and polygenic obesity. Monogenic obesity is a rare and severe condition that typically manifests early in life and is attributed to a single pathogenic variant in one of several genes—an inventory that continues to grow^8,9^. In contrast, polygenic obesity is more common and arises from the aggregate effect of multiple genetic variants at different loci, each contributing modestly to overall risk^10^. Gene panels used to assess monogenic obesity can contain up to 57 loci such as that used in the study of Kunzel et al.^9^. Main genes causing severe phenotypes of monogenic obesity seem to be from key neuroendocrine circuits, predominantly involving the leptin-melanocortin pathway, such as *LEP, LEPR, POMC, MC4R, PCSK1, SIM1, NTRK2, SH2B1* and *BBS* genes^11^. Hundreds of genes are associated with polygenic obesity with small contributions from each such genes; the most-often discussed genes for polygenic obesity include FTO, MC4R, *SEC16B, LINC02702, GNPDA2, BDNF, ADCY3, RALGAPA1, IRX3/IRX5, and ZNF259* ^11^. Recent findings suggest that monogenic and polygenic obesity share underlying biological mechanisms, with the brain playing a pivotal role in regulating body weight^12^; common neuroendocrine signaling pathways that primarily affect the central regulation of appetite and energy balance of the brain exists between both the forms of obesity^11^.

Numerous genome-wide association studies (GWAS) have identified several ‘common’ genetic variants associated with polygenic obesity traits, mainly body mass index (BMI). Polygenic score (PGS) models for obesity risk built using such GWAS BMI-associated common variants are advocated for use in obesity prognosis to initiate timely intervention. However, the latest GWAS BMI-based PGS models could explain only 7.2% to 17.5%, depending on ethnicity, of the variation in BMI^13^. Earlier models explained even less variation and their ability to accurately predict the risk of adult obesity remains modest (AUC_ROC_ ∼0.64)^10^. The role of rare and low-frequency genetic variants in the susceptibility of an individual to polygenic obesity is increasingly recognized in recent literature: **(i)** Turcot et al.^14^ reported 14 rare and low-frequency coding variants, of which 8 were in genes newly implicated in obesity (*ZBTB7B, ACHE, RAPGEF3, RAB21, ZFHX3, ENTPD6, ZFR2* and *ZNF169*), 2 in genes previously observed to be mutated in extreme obesity (*MC4R* and *KSR2*), and 2 in *GIPR*. Notably, the BMI-increasing variants from this list were also associated with an elevated risk of weight regain after bariatric surgery in adults of European ancestry with morbid obesity^15^; **(ii)** Akbari et al.^16^, analysed whole-exomes of ∼640,000 individuals from the UK, the US, and Mexico, identifying low-frequency protein-coding variants from 16 genes associated with BMI; **(iii)** Jiao et al. identified low-frequency coding variants associated with morbid obesity from the *SYPL2*^17^ and *LAMB3* genes^18^ using exome sequencing in Swedish individuals.

Ethnic background contributes substantially to differences in the prevalence and genetic architecture of obesity. The Middle East region exemplifies this diversity, exhibiting a distinct and a notably high obesity profile. According to the World Health Organization 2022 data on the prevalence of obesity among adults (https://www.who.int/data/gho), Kuwait and Qatar ranked 10^th^ and 11^th^ globally, with obesity rates of 45.2% and 43.8%, respectively. The genetic make-up of the region is unique as it has traditionally practised consanguinity and endogamy resulting in increased homozygosity and accumulation of recessive alleles. The potential contribution of consanguinity to polygenic obesity has been previously highlighted^19,20^. The rapid shifts in lifestyle and diet-patterns experienced in the oil-rich era, combined with the long tradition of consanguinity, have conferred a unique genetic and clinical susceptibility profile of obesity to the region; studying such a population may uncover additional pathways underlying obesity. A recent systemic review of obesity genetics studies from Arab countries^21^ revealed two variants unique to Arab populations and 19 variants distinctively associated with obesity in Arabs that had previously been associated with other clinical conditions in non-Arabs.

Given the increasing health burden posed by obesity—particularly morbid obesity—this study investigates the spectrum of rare and low-frequency genetic risk variants associated with morbid obesity among Arab adults from Kuwait and Qatar. The objectives were to: (1) characterize rare and low-frequency protein-coding deleterious genetic variants implicated in polygenic morbid obesity within this population; and (2) evaluate the cumulative impact of these variants on BMI across overweight, obese, and morbidly obese groups.

## Results

### Clinical characteristics of the study participants

The study considered two cohorts – 285 native Kuwaiti adults (67 morbidly obese, 85 obese, 65 overweight, and 68 normal-weight individuals), and 853 native Qatari adults (123 morbidly obese, 403 obese, 243 overweight, and 86 normal-weight individuals). Participants were generally healthy, except for individuals with T2D or hypertension. Pregnant women, and individuals with rare genetic disorders or chronic diseases were excluded. The demographic and clinical characteristics of the study cohorts are summarized in **Table 1**. The mean BMI of morbidly obese individuals were 46±5 kg/m^2^ (Kuwait) and 45±5 kg/m^2^ (Qatar). Among Kuwaiti participants, morbidly obese individuals exhibited a higher mean age compared to normal-weight individuals (56±17 versus 38±15 years). Age-matching was more consistent in the cohort from Qatar: mean age of morbidly obese individuals was 50±10 years while that of normal-weight individuals was 47±12 years. Among Kuwaiti participants, the morbidly obese category exhibited a higher proportion of females (72%) than the normal-weight category (56%). Conversely, among Qatari participants, females were more prevalent in the normal-weight group (87%) compared to the morbidly obese group (55%). The prevalence rates of T2D and hypertension in the Kuwaiti cohort were significantly elevated in the morbidly obese category compared to the normal-weight category (T2D: 61% versus 19%; Hypertension: 58% versus 5.9%). In contrast, among the Qatari cohort, the prevalence of T2D showed minimal difference between morbidly obese and normal-weight groups (47% versus 41%).

**Table 1.** Summary statistics of the study cohorts from Kuwait and Qatar.

| **1.a. Study Cohorts from Kuwait.** |  |  |  |  |
| --- | --- | --- | --- | --- |
| **Variable** | **Control N = 68<sup>1</sup>** | **Overweight/Obese N = 150<sup>1</sup>** | **Morbid obese N =** | **p-value<sup>2</sup>** |
| Age (years) | 38 ± 15 | 52 ± 15 | 56 ± 17 | <0.001 |
| Sex |  |  |  | 0.073 |
| F | 38 (56%) | 84 (56%) | 48 (72%) |  |
| M | 30 (44%) | 66 (44%) | 19 (28%) |  |
| BMI (kg/m<sup>2</sup>) | 23 ± 1 | 31 ± 4 | 46 ± 5 | <0.001 |
| Waist Circumference (cm) | 78 ± 8 | 100 ± 12 | 126 ± 13 | <0.001 |
| Type 2 diabetes | 13 (19%) | 69 (46%) | 41 (61%) | <0.001 |
| Hypertension | 4 (5.9%) | 61 (41%) | 39 (58%) | <0.001 |
| Variants per sample (Median (IQR)) from KU_374 variant | NA | 4.5 (3.0, 7.0) | 12.0 (9.0, 16.0) | <0.001 |
| **1.b. Study Cohorts from Qatar.** |  |  |  |  |
| **Variable** | **Control N = 86<sup>1</sup>** | **Overweight/Obese N = 644<sup>1</sup>** | **Morbid obese N =** | **p-value<sup>2</sup>** |
| Age (years) | 47 ± 12 | 54 ± 11 | 50 ± 10 | <0.001 |
| Sex |  |  |  | <0.001 |
| Female | 75 (87%) | 361 (56%) | 68 (55%) |  |
| Male | 11 (13%) | 283 (44%) | 55 (45%) |  |
| BMI (kg/m<sup>2</sup>) | 23 ± 2 | 32 ± 4 | 45 ± 5 | <0.001 |
| Type 2 diabetes | 35 (41%) | 398 (62%) | 58 (47%) | <0.001 |
| Variants per sample (Median (IQR)) from QA_1155 variant |  | 11 (8, 14) | 23 (18, 28) | <0.001 |

<sup>1</sup>Mean ± SD; n (%); <sup>2</sup>Kruskal-Wallis rank sum test; Pearson's Chi-squared test.

### Rare and low-frequency deleterious genetic risk variants associated with morbid obesity

Whole-exome sequencing generated ∼100 million non-duplicated sequencing reads per individual. The mean sequencing coverages were 44X and 57X in Kuwaiti and Qatari cohorts, respectively. On an average, at least a 15X coverage was observed for ∼80% of the targeted base pairs. Mapping the sequencing reads to the human reference genome (hg19/GRCh37) yielded an initial set of 168,898 and 374,264 variants in the Kuwaiti and Qatari cohorts, respectively. Comprehensive variant analysis (see Methods and **Supplementary Fig. 1**) narrowed these to 374 rare and low-frequency deleterious variants from 356 genes and 1155 from 1065 genes differentiating morbidly obese individuals from normal-weight individuals in the Kuwaiti and Qatari cohorts, respectively (see **Supplementary Datasets D1** and **D2** for the KU_374 and QA_1155 variants lists). Density maps and scatter plots of minor allele frequency (MAF) distributions of these variants (**Supplementary Fig. 2)** indicated skewed distributions toward an allele frequency below 0.015 in both the regional dataset (Greater Middle Eastern (GME) Variome database – Arabian Peninsula subset^22^) and the global dataset (Genome Aggregation Database *-* gnomAD_all Genomes v2.1.1^23^).

### Distribution and burden of morbid obesity risk variants from Kuwait

Each of the KU_374 morbid obesity risk variants were present in at least 2 Kuwaiti morbidly obese individuals and absent in all normal-weight individuals. All the 67 Kuwaiti morbidly obese individuals harboured one or more of these variants. Furthermore, 94% of the obese individuals and 74% of the overweight individuals carried at least one of these variants (**Table 2A**). Increasing BMI across the Kuwaiti overweight, obese, and morbidly obese categories showed a positive correlation (r=0.63, 95% CI [0.46, 0.76]; p<0.001) with the count of such harboured variants (**Fig. 1A**, **Table 3**). The density plots of variant count by BMI indicated that the density peaked at a higher count of variants in the case of morbidly obese category compared to the overweight and obese categories (**Fig. 2A**). The median (IQR) of variants counts per individual in the Kuwaiti overweight, obese, and the morbidly obese categories were 5 (3.0, 7.0), 4 (3.0, 6.0) and 12.0 (9.0, 15.0), respectively (**Table 2B**).

**Fig. 1.**
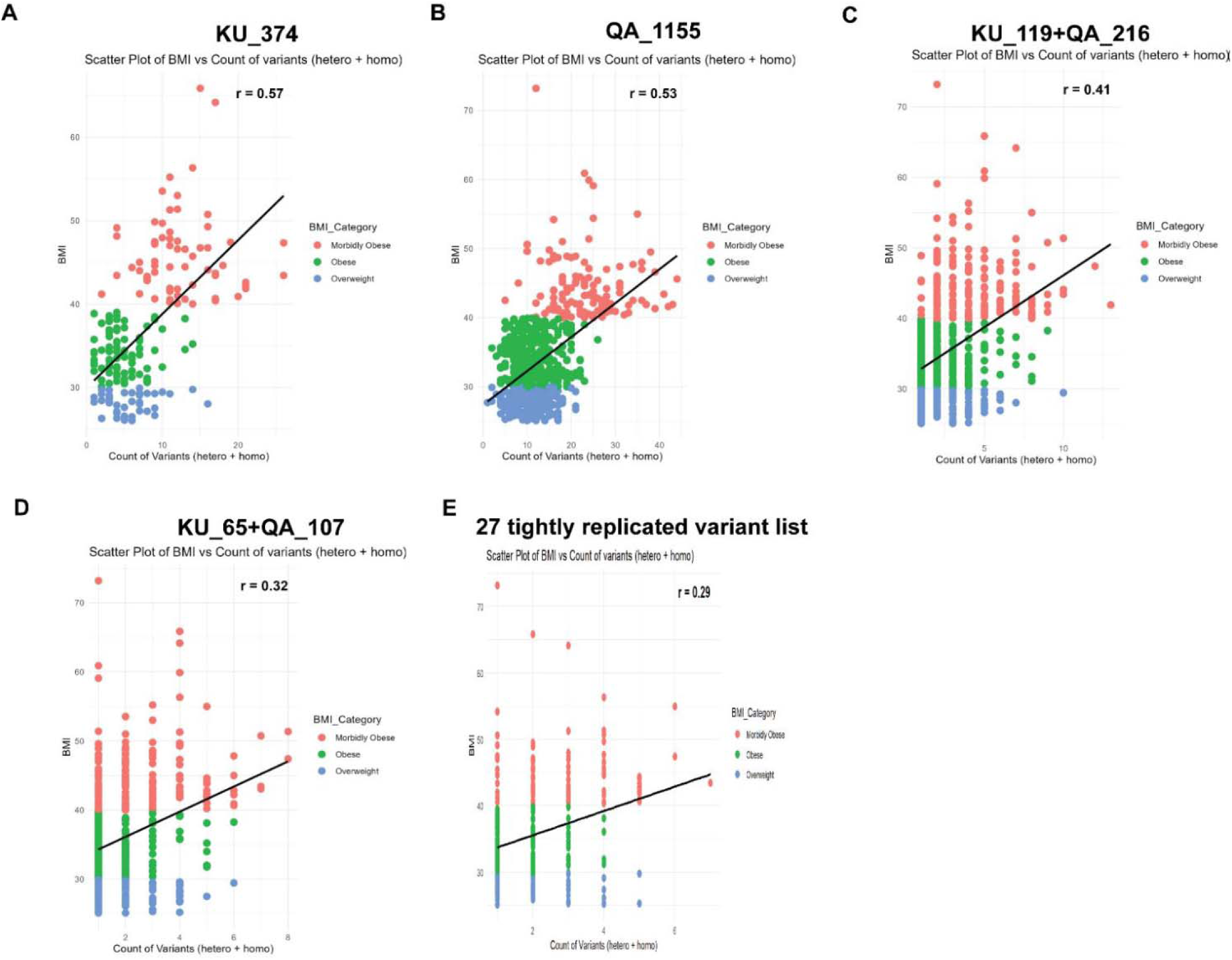
Scatter plot of BMI with count of variants. (A) using the KU_374 variants on the Kuwaiti sample set of overweight, obese and morbidly obese individuals; (B) using the QA_1155 variants on the Qatari cohorts; (C) using the loosely-replicated KU_119+QA_216 variants on the combined cohorts; (D) using the leniently-replicated KU_65+QA_107 variants on the combined cohorts; and (E) using the tightly-replicated 27 variants on the combined cohorts.

**Fig. 2.**
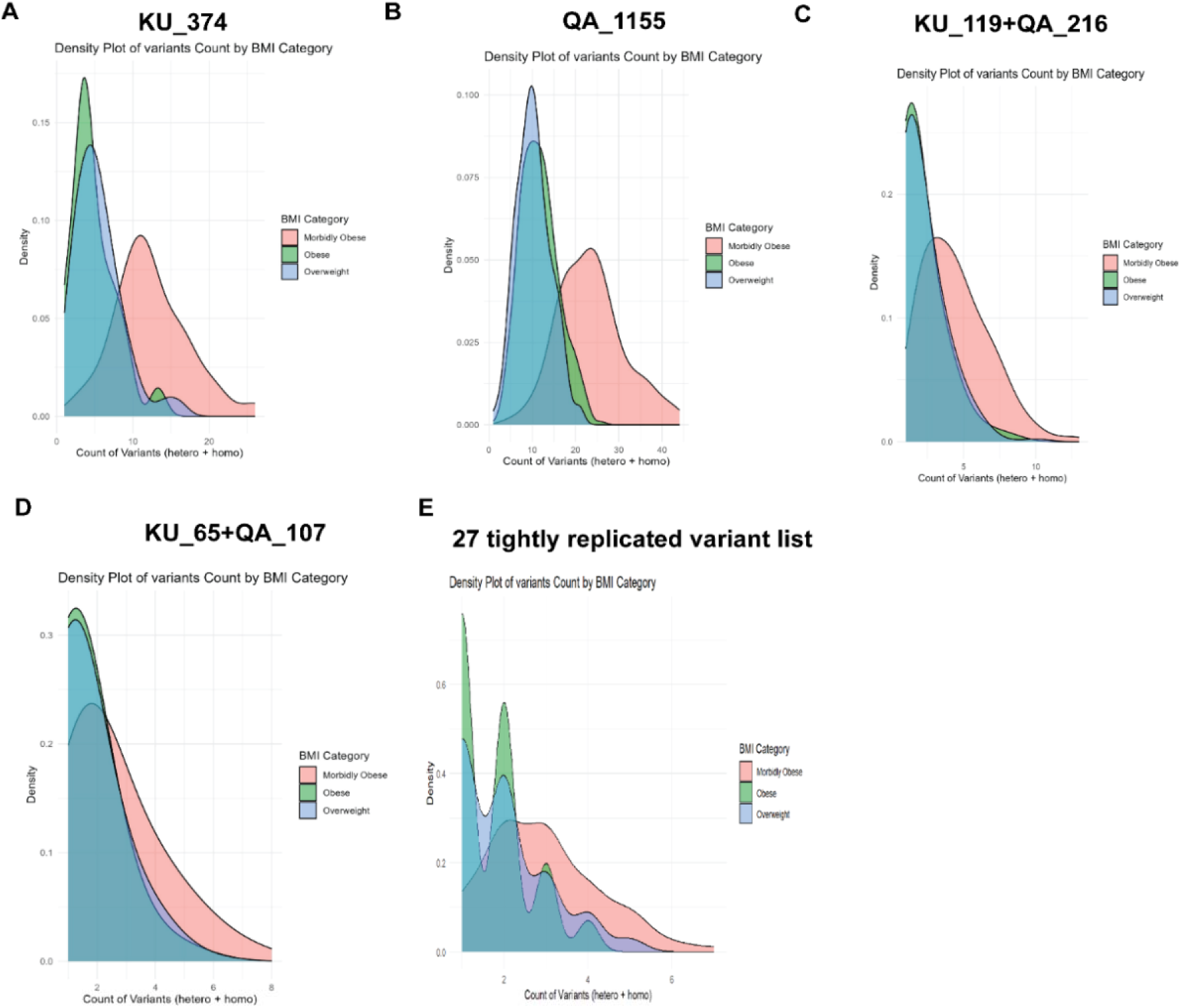
Density map of BMI with count of variants. (A) using the KU_374 variants on the Kuwaiti sample set of overweight, obese and morbidly obese individuals; (B) using the QA_1155 variants on the Qatari sample set of overweight, obese and morbidly obese individuals; (C) using the loosely-replicated KU_119+QA_216 variants on the merged sample set of the two cohorts; (D) using the leniently-replicated KU_65+QA_107 variants on the merged sample set of the two cohorts; and (E) using the tightly-replicated 27 variants list on the merged sample set of the two cohorts.

**Table 2:**
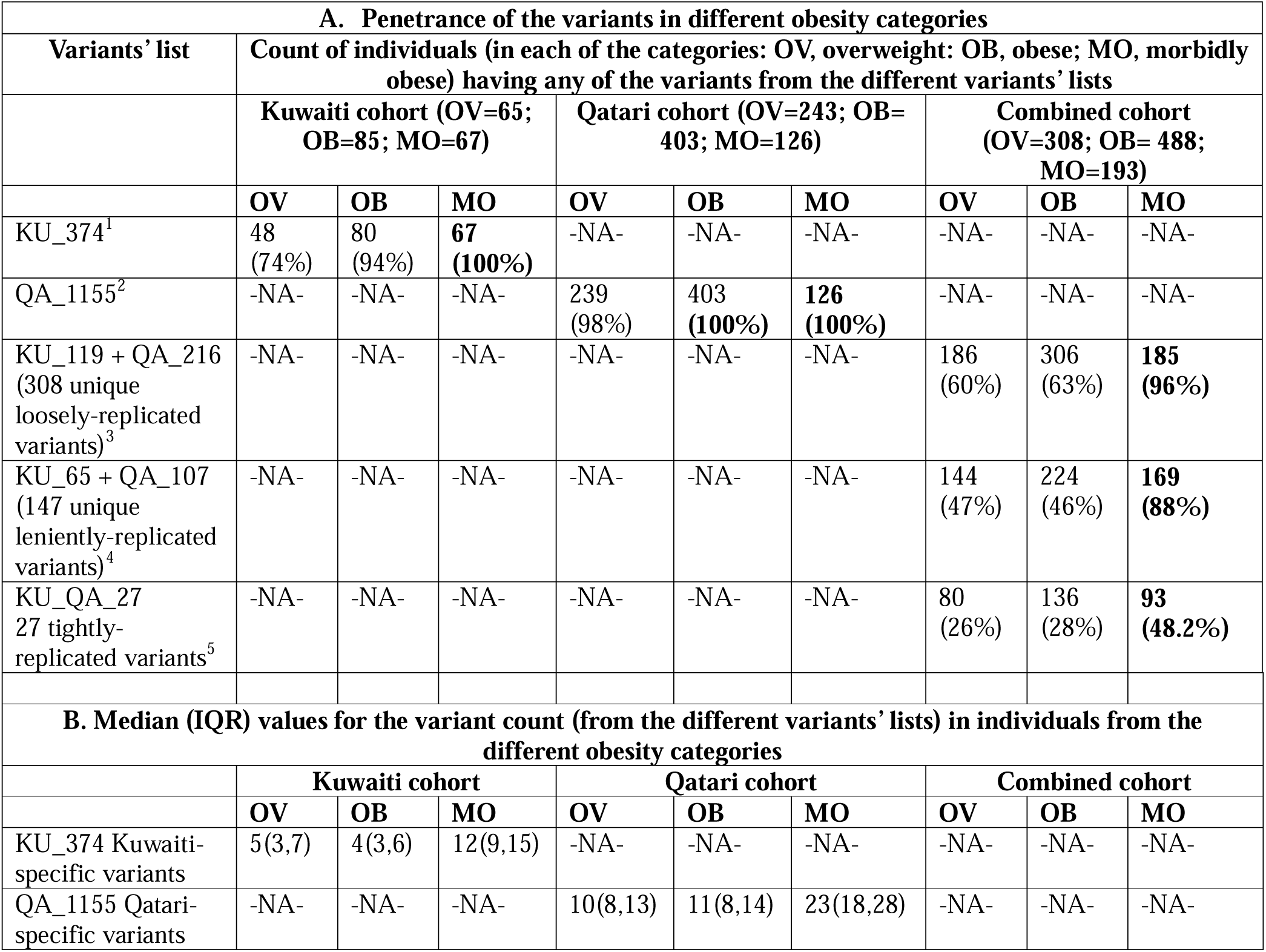
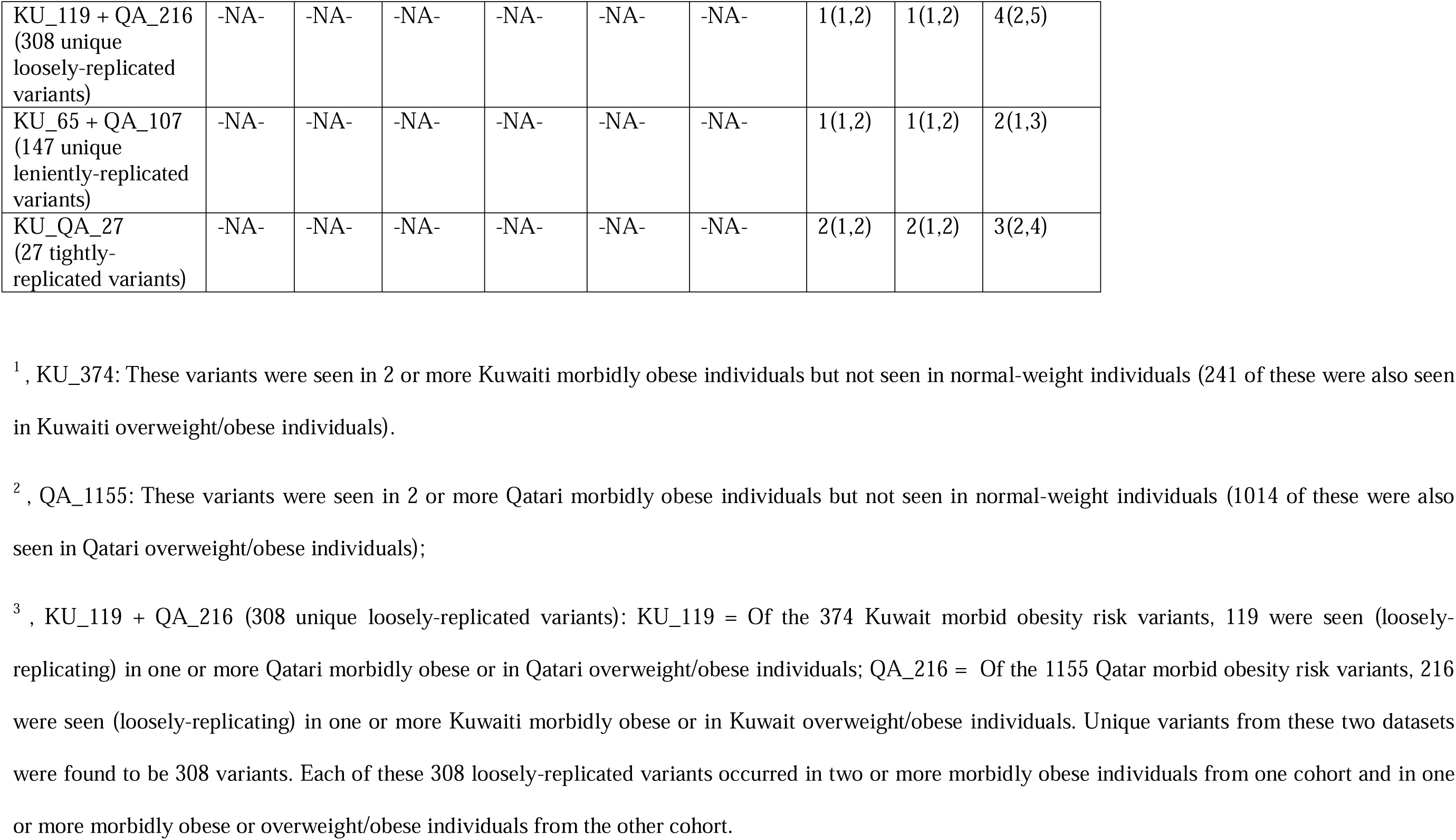
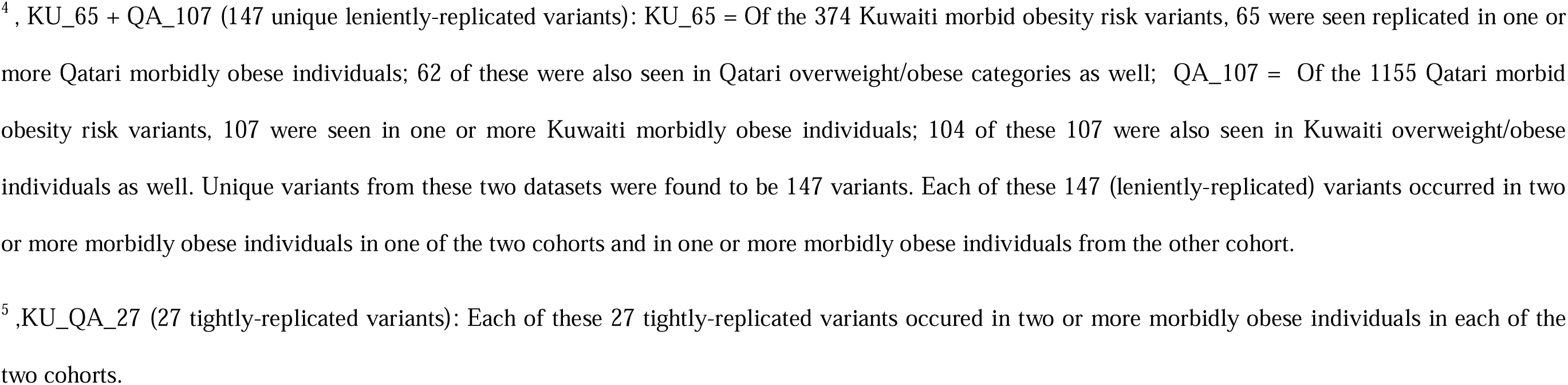
Penetrance and burden of variants from the different variants’ lists generated in the study.

**Table 3.** BMI correlations with variant burden for the different variants’ lists generated in the study.

| Variants' list<sup>1</sup> | Count of unique variants and genes | Count of genes having GWAS annotation for obesity traits | BMI correlation statistics (r and 95% CI) with p<0.001 in |  |  |
| --- | --- | --- | --- | --- | --- |
|  |  |  | Kuwaiti cohort | Qatari cohort | Combined cohort |
| KU_374 | 374/356 | 201 (56.5%) | **0.63, 95% CI [0.46, 0.76]** | -NA- | -NA- |
| QA_1155 | 1155/1064 | 685 (64.4%) | -NA- | **0.57, 95% CI [0.47, 0.65]** | -NA- |
| KU_119 + QA_216 (308 unique **loosely-replicated variants**) | 308/288 | 180 (62.5%) | -NA- | -NA- | **0.41, 95% CI [0.31, 0.50]** |
| KU_65 + QA_107 (147 unique **leniently-replicated variants**) | 147/142 | 88 (62%) | -NA- | -NA- | **0.30, 95% CI [0.20, 0.40]** |
| KU_QA_27<br>27 **tightly-replicated variants** | 27/27 | 13 (48%)<sup>2</sup> | -NA- | -NA- | **0.31, 95% CI [0.20, 0.41]** |

<sup>1</sup>, for footnotes on variants list, refer to Table 2.<sup>2</sup>, see Table 4 for detailed annotation from GWAS Catalog for each of the 27 gene loci.

### Distribution and burden of morbid obesity risk variants from Qatar

Each of the QA_1155 morbid obesity risk variants were present in at least 2 Qatari morbidly obese individuals but not in any of the normal-weight individuals. All the 126 morbidly obese and the 403 obese individuals from Qatar harboured one or more of these variants. Furthermore, only 4 of the 243 overweight individuals did not exhibit any of these variants (**Table 2A**). Increasing BMI across the Qatari overweight, obese and morbidly obese categories showed a positive correlation (r=0.57, 95% CI [0.47, 0.65]; p<0.001) with the number of such harboured variants (**Fig. 1B**, **Table 3**). The density plots of variant count by BMI indicated that the density peaked at a higher count of variants in the case of morbidly obese category compared to the overweight and obese categories (**Fig. 2B**). The median (IQR) of variants counts per individual in the Qatari overweight, obese and the morbidly obese categories were 10.0 (8.0, 13.0), 11.0 (8.0, 14.0) and 23.0 (18.0, 28.0), respectively (**Table 2B**).

### Replicated variants between the Kuwaiti and Qatari cohorts and the variant burden on BMI levels in the combined cohort of Kuwaiti and Qatari participants

#### Loosely-replicating variants

119 of the KU_374 variants were seen loosely-replicating in one or more Qatari morbidly obese and/or overweight/obese individuals; and 216 of QA_1155 were seen loosely-replicating in one or more Kuwaiti morbidly obese and/or overweight/obese individuals (**Supplementary Fig. 3**). Combining the loosely-replicating variants from these two sets resulted in 308 variants (KU_119+QA_216 list) from 288 genes (**Supplementary Dataset D3**). Only 8 of the 193 morbidly obese individuals from the combined cohort of Kuwait and Qatar did not exhibit any of these variants (**Table 2A**). Up to 60% of the overweight individuals and 63% of the obese individuals carried one or more of these loosely-replicated variants. Across the combined cohort, increasing BMI among overweight, obese and morbidly obese categories was positively associated with the count of harboured variants (r=0.41, 95% CI [0.31, 0.50]; p<0.001) (**Fig. 1C**, **Table 3**). Density plots of variant count by BMI category revealed a rightward shift in the distribution peak for morbidly obese category compared to the overweight or obese categories, indicating a higher variant burden among morbidly obese participants **(Fig. 2C)**. The median (IQR) number of loosely-replicated variants per individual in the overweight, obese and the morbidly obese categories were 1.0 (1.0, 2.0), 1.0 (1.0, 2.0) and 4.0 (2.0, 5.0), respectively (**Table 2B**).

#### Leniently-replicated-variants

Requiring that a morbid obesity risk variant in one cohort be present in one or more of the morbidly obese individuals from the other cohort (irrespective of its occurrence in overweight/obese individuals) reduced the count of Kuwait-to-Qatar loosely-replicated variants to 65 leniently-replicated variants and the Qatar-to-Kuwait loosely-replicated variants to 107 leniently-replicated variants (**Supplementary Fig. 3**). The combined set of these two sets of leniently-replicated variants resulted in 147 KU_65+QA_107 variants across 142 genes (**Supplementary Dataset D4**). 169 (88%) of the 193 morbidly obese individuals from the combined cohort carried one or more of these 147 leniently-replicated variants; and 46% of the obese and 47% of the overweight individuals harboured at least one of these 147 variants (**Table 2A**). Variant count correlated positively with BMI (r=0.30, 95% CI [0.20, 0.40]; p<0.001) **(Fig. 1D**, **Table 3).** Density plot of variant count by BMI showed an increasing burden with higher BMI categories **(Fig. 2D). T**he median (IQR) values for these variants counts per individual were 1.0 (1.0, 2.0) in the overweight/obese categories and 2.0 (1.0, 3.0) in the morbidly obese category (**Table 2B**).

#### Tightly-replicated morbid obesity risk variants (prioritised candidate variants)

Upon applying a stricter replication criterion requiring that each variant is present in at least two morbidly obese individuals from each of the two cohorts (**Supplementary Fig**. **3**), the list of leniently-replicated variants narrowed down to a set of 27 tightly-replicated variants (**Table 4)**. Among the 193 morbidly obese individuals from the combined cohort, 93 (48.2%) harboured one or more of these 27 variants. Additionally, 26% of the overweight and 28% of the obese individuals carried at least one of these variants (**Table 2A**). Analysis of the association between the burden of these 27 variants and BMI across the overweight, obese, and morbidly obese individuals revealed a statistically significant correlation coefficient of r=0.31, 95% CI [0.20, 0.41]; p<0.001) (see **Fig. 1E**, **Table 3**). Mean values for the count of these 27 variants were 1.96±1.05 in the overweight group, 1.74±0.85 in the obese group, and 2.89±1.31 in the morbidly obese group. The density plot showed a rightward shift in the peak for morbidly obese individuals, as presented in **Fig. 2E**.

**Table 4.**
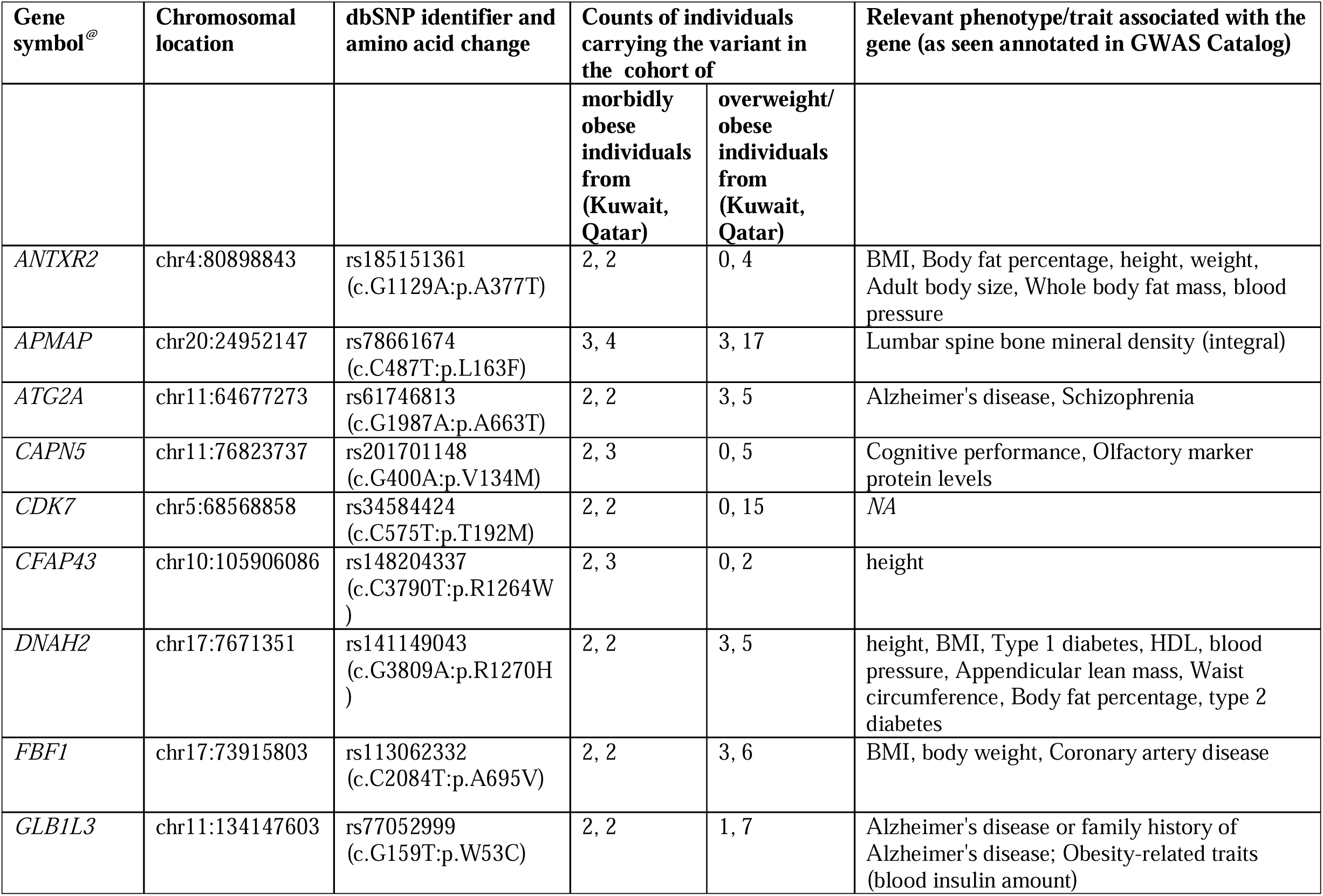
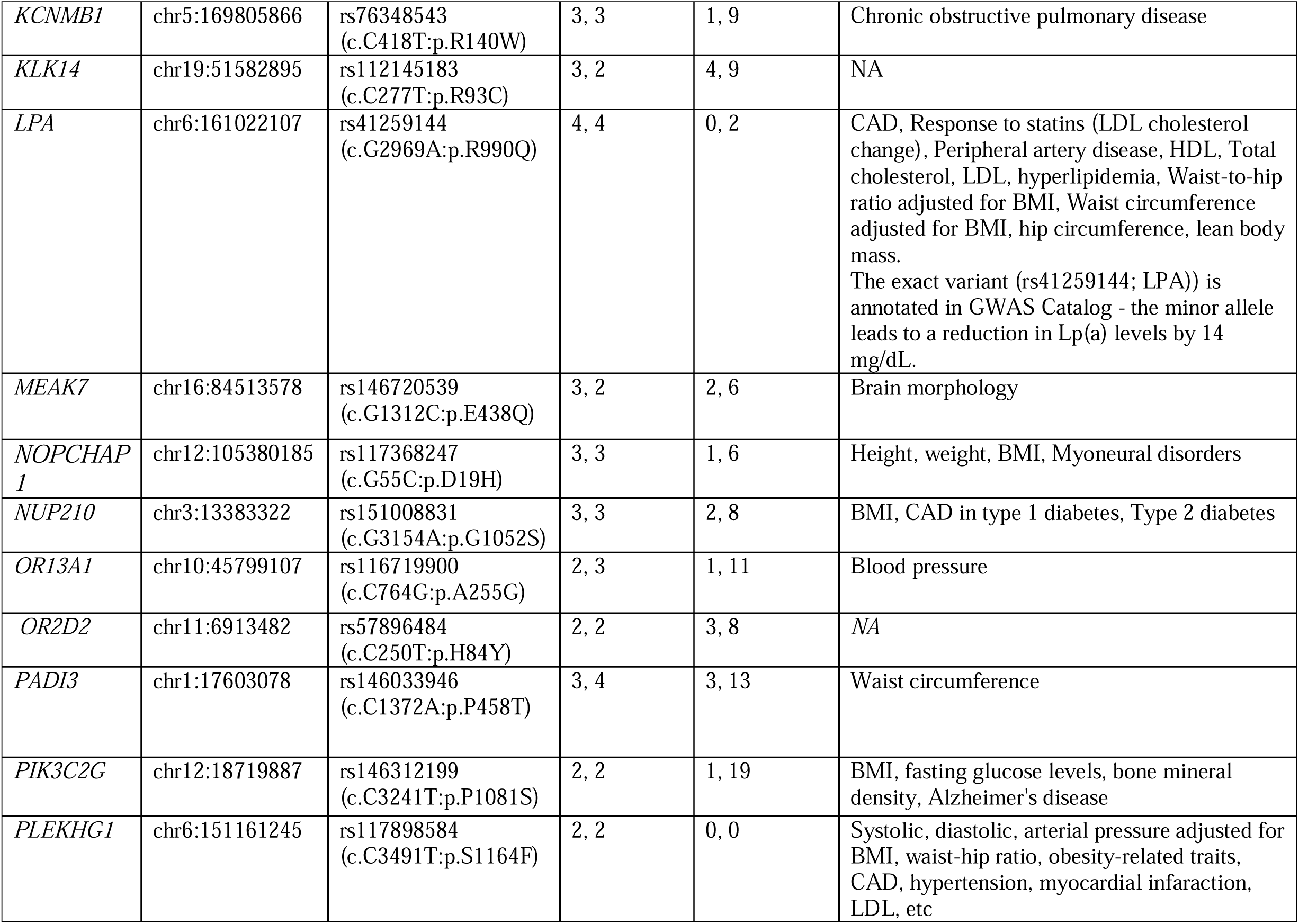
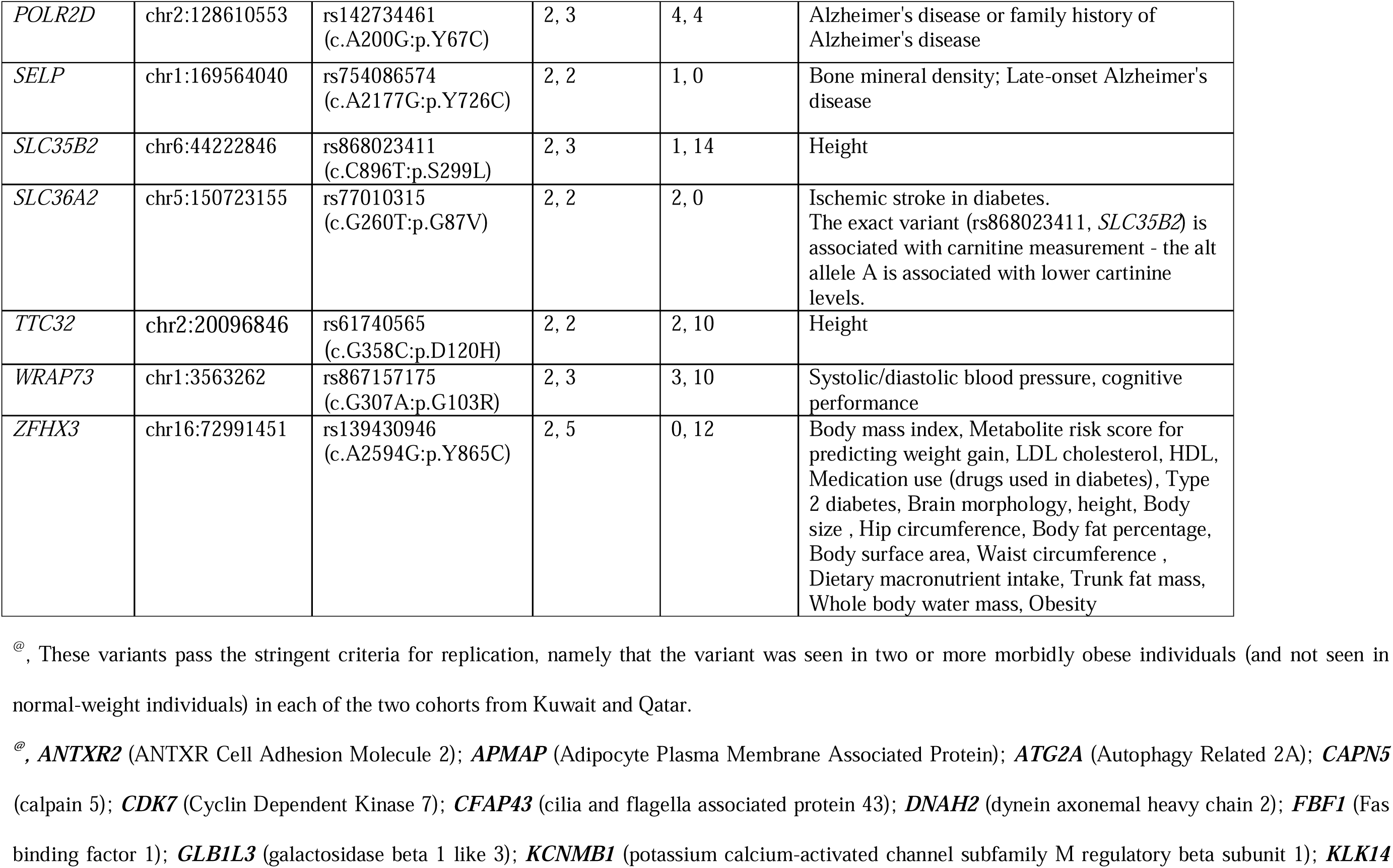
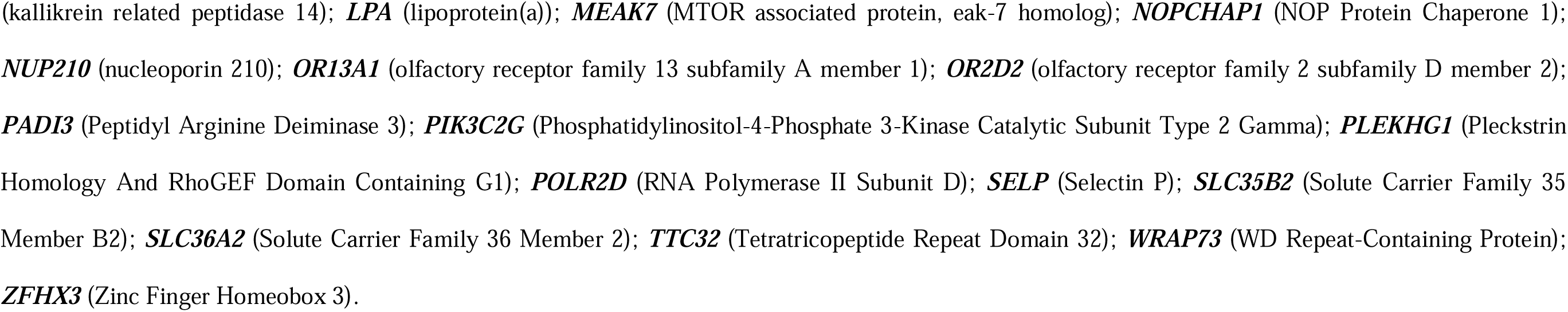
List of the 27 tightly-replicated morbid obesity risk variants^&^.

### Pathway enrichment and tissue-specific expression enrichment analyses

Pathway enrichment analyses of the morbid obesity risk gene loci from the derived variants’ lists are illustrated in **Fig. 3A-E**. Top-enriched pathways included terms reflecting metabolic and adipogenic processes underlying obesity – such as white adipose ass & tissue mmorphology, fat cell morphology, increased total body fat, increased epididymal fat pad amount, increased body mass, decreased lean body mass, lipid metabolism, insulin secretion/resistance, and mTOR signaling.. Key enriched tissues that resulted from expression data analysis using GTEx^24^ reflected a predominant role for adipose and metabolic tissues in obesity pathophysiology (**Fig. 4A-E**). Key tissues enriched for expression by the morbid obesity risk gene loci from the derived variants’ lists include adipose tissue, smooth muscle, skeletal muscle, whole blood, liver, and and pancreas. The identified risk genes such as *LPA*, *SLC36A2*, *PIK3C2G*, and *APMAP* showed preferential expression in metabolic tissues, supporting their potential roles in obesity pathophysiology.

**Fig. 3.**
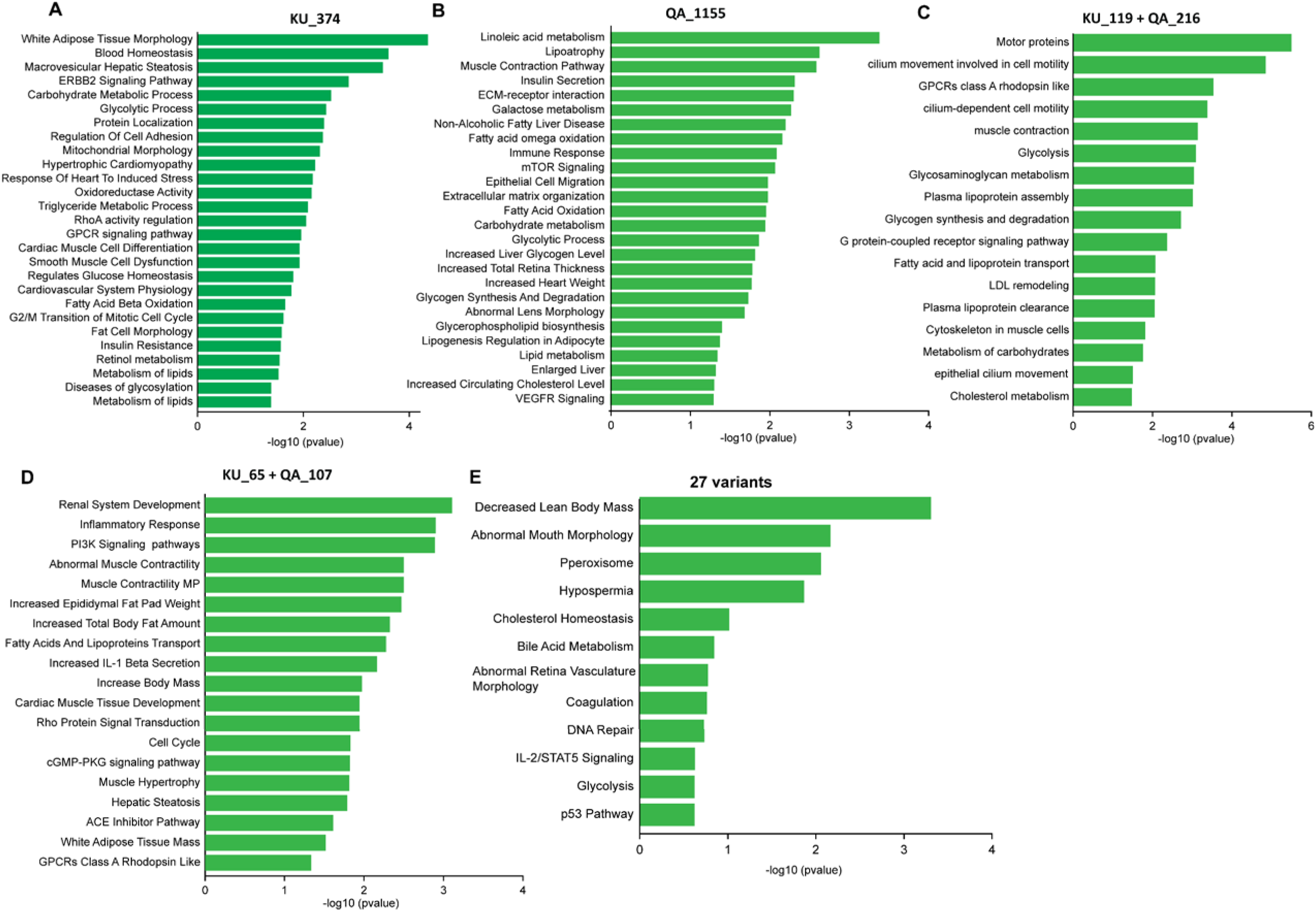
Pathway enrichment analyses of the morbid obesity risk gene locis identified in Kuwaiti and Qatari cohorts. Bar plots display significantly enriched biological pathways associated with the risk gene loci identified from the exome-wide burden analyses. **(A) Enriched pathways derived using the morbid obesity risk gene loci from the Kuwaiti cohort (KU_374 variants’ list):** Top pathways include white adipose tissue morphology, fat cell morphology, insulin resistance, and metabolism of lipids (and abnormal adipose tissue physiology, increased body fat amount, and abnormal glucose homeostasis), reflecting metabolic and adipogenic processes underlying obesity. **(B) Enriched pathways derived using the morbid obesity risk gene loci from the Qatari cohort (QA_1155 variants’ list):** Major enriched terms include insulin secretion, NAFLD, mTOR signaling, fatty acid oxidation, lipid metabolism, and cholesterol levels (and increased body mass, increased epididymal fat pad weight, white adipose tissue mass, and abnormal lipid homeostasis), reflecting an adipose-centric obesity mechanism. **(C)** Enriched pathways derived using the morbid obesity risk gene loci from the loosely-replicated set of KU_119 + QA_216 variants’ list: Prominent terms include fatty acid and lipoprotein transport, LDL remodeling, and cholesterol metabolism, (and increased total body fat amount, abnormal adipose tissue physiology, decreased brown adipose tissue amount, and abnormal glucose tolerance), representing convergent molecular signatures across the two cohorts. **(D) Enriched pathways derived using the morbid obesity risk gene loci from the leniently-replicated set of KU_65 + QA_107 variants’ list:** Enriched pathways from the leniently-replicated set of 147 variants seen exclusively in morbidly obese individuals in both the cohorts. Key pathways include increased epididymal fat pad amount, increased total body fat, increased body mass, white adipose tissue mass, and cardiac muscle tissue morphology, (and fat cell morphology, abnormal insulin sensitivity) highlighting cross-cohort conserved molecular disruptions. **(E) Pathway enrichment analysis for the morbid obesity risk genes harboring the 27 tightly-replicated variants**: Key pathways include decreased lean body mass, and cholesterol homeostasis, IL-2/STATS signaling, and p53 pathway.

**Fig. 4.**
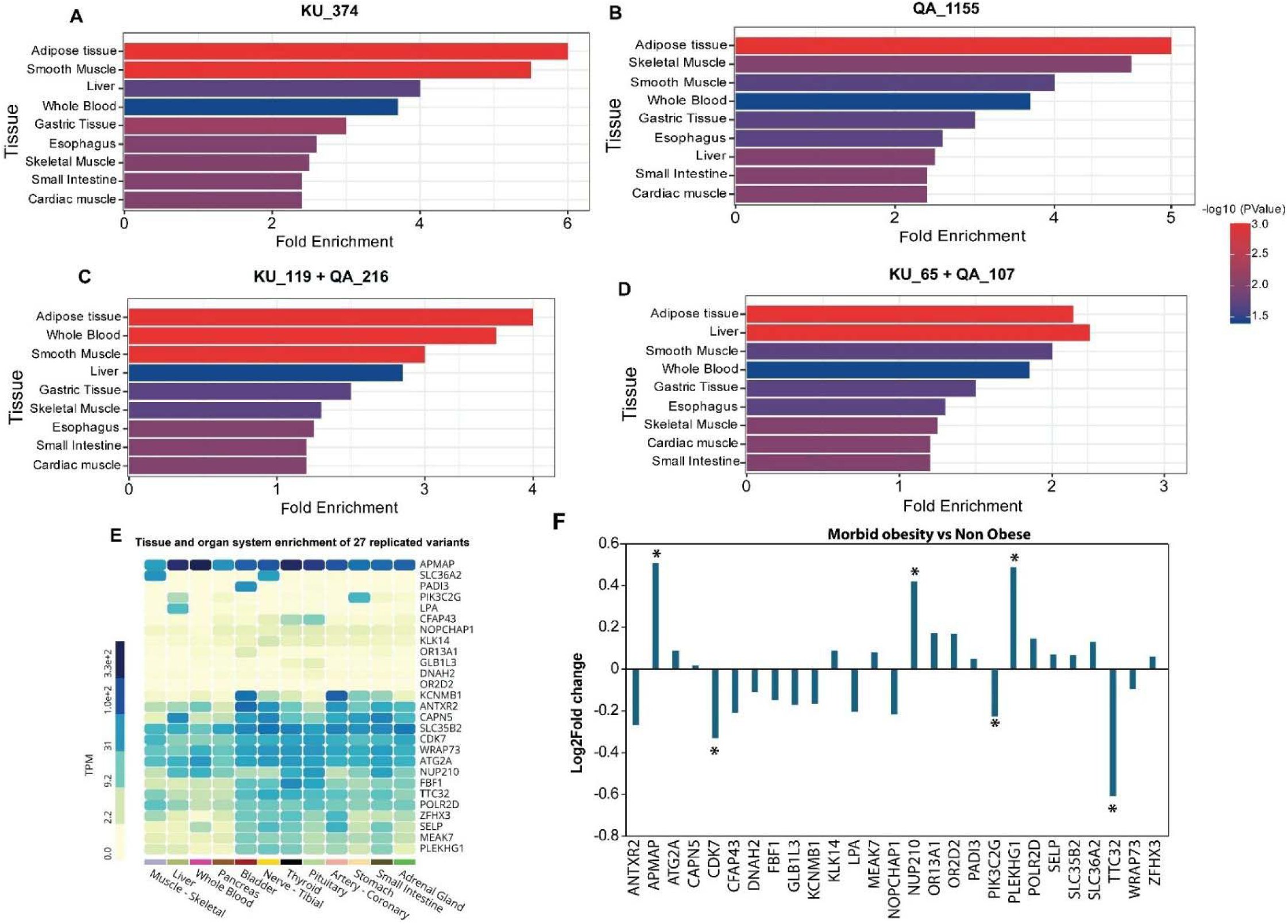
Gene-based tissue-specific expression enrichment analysis for the identified morbid obesity risk gene loci using GTEx or GSE147562 GEO dataset. The bar plots display the tissues significantly enriched for expression of the risk genes harboring the rare and low-frequency variants identified in Kuwaiti, Qatari, and combined cohorts. The color gradient represents the –log (p-value) of enrichment, and the X-axis indicates fold enrichment of expression across tissues. (A) Tissue enrichment for morbid obesity risk gene loci from the Kuwaiti cohort (KU_374 variants’ list): Adipose tissue, smooth muscle, and liver exhibited the strongest enrichment, reflecting a predominant role of adipose and metabolic tissues in obesity pathophysiology. (B) Tissue enrichment for variants from the Qatari cohort (QA_1155 variants’ list): Adipose, skeletal, and smooth muscle tissues were the most enriched. (**C**) **Tissue enrichment for the loosely-replicated variants across Kuwaiti and Qatari cohorts (KU_119 + QA_216 variants’ list)**: Adipose tissue and whole blood showed the highest enrichment, suggesting shared systemic metabolic and inflammatory involvement across populations. (**D**) **Tissue enrichment for leniently-replicated morbid obesity variants across the Kuwaiti and Qatari cohorts (KU_65 + QA_107 variants’ list):** Adipose, liver, and smooth muscle tissues were the most enriched. (**E**) **GTEx-based tissue and organ system expression heatmap showing transcripts per million (TPM) values for the 27 tightly-replicated morbid obesity risk gene loci across human tissues:** High expression was observed in adipose tissue, skeletal muscle, liver, and pancreas, emphasizing their metabolic importance. Genes such as *LPA*, *SLC36A2*, *PIK3C2G*, and *APMAP* showed preferential expression in metabolic tissues, supporting their potential roles in obesity pathophysiology. **(F) Differential expression analysis of the 27 tightly-replicated risk genes in duodenal tissue from morbidly obese individuals *versus* non-obese normal-weight individuals using the GSE147562 dataset:** The identified tightly-replicated morbid obesity risk genes, namely *APMAP*, *CDK7*, *NUP210*, *PLEKHG1* and *TTC32,* that showed significant changes in expression (p-value ≤ 0.05) between morbidly obese and non-obese individuals are marked with asterisks. The genes from the leniently-replicated set of risk genes, that are differentially expressed in morbidly obese individuals are presented in **Supplementary Table 2.**

### Analysing the identified risk loci for differential expression in morbidly obese individuals using publicly available datasets

Expression analysis of the risk genes, using the publicly available transcriptional profile of duodenal tissue from morbidly obese individuals and non-obese individuals generated on a Spanish cohort by Ho-Plágaro et al. ^25^ (Gene Expression Omnibus (GEO) identifier: GSE147562), revealed that six of the 27 tightly-replicated risk genes (namely *APMAP, CDK17, NUP210, PIK3C2G, PLEKHG1* and *TTC32*) exhibited significant dysregulation (p-value ≤ 0.05) in morbidly obese individuals compared to normal-weight individuals (**Fig. 4F; Supplementary Table 1)**; the *APMAP* and the *TTC32* exhibited a log2 fold change (log2FC) of ≥ 0.5. Among the 142 risk genes corresponding to the 147 leniently-replicated risk variants, 27 genes were significantly dysregulated (p-value ≤ 0.05) in morbidly obese individuals compared to normal-weight individuals; of these, the *MPEG1, DOCK2, SEMA4D, ZNF280D* (along with the *APMAP* and *TTC32*) genes exhibited a log2FC value of ≥ 0.5 (**Supplementary Table 2**).

### Replicated variants characterising the top nine BMI-ranked morbidly obese individuals

Examination of the top nine BMI-ranked individuals for harbouring the replicated variants (**Table 5**) revealed that five individuals, with a BMI of 73.2, 65.9, 64.17, 56.32, and 55 kg/m^2^, harboured five of the 27 tightly-replicated variants – the *SELP* variant (in individuals with BMI of 73.2, 65.9, and 56.32 kg/m^2^), *PADI3* variant (in the individual with a BMI of 65.9 kg/m^2^}, *OR2D2* variant (in the individual with a BMI of 64.17 kg/m^2^), the *NOPCHAP1* variant (in the individual with a BMI of 56.32 kg/m^2^), and the *GLB1L3 variant* (in the individual with a BMI of 55.0 kg/m^2^). Upon performing genetic association tests for these variants with BMI as continuous variable among morbidly obese individuals from the combined cohort, we found the effect sizes to be significantly large for three of them; the SELP/rs754086574 variant exhibited an effect size of 27.41.(standard error: 8.4; p-value=0.001), the PADI3/rs146033946 variant exhibited an effect size of 17.33.(standard error: 7.0; p-value=0.014); and the OR2D2/rs57896484 variant exhibited an effect size of 20.35.(standard error: 8.5; p-value=0.018). Certain replicated variants enrich these top nine BMI-ranked morbidly obese individuals – *SELP*-rs754086574 in three individuals, and each of *EPPK1*-rs150047119 and *MCM4*-rs149232439 in two individuals.

**Table 5.**
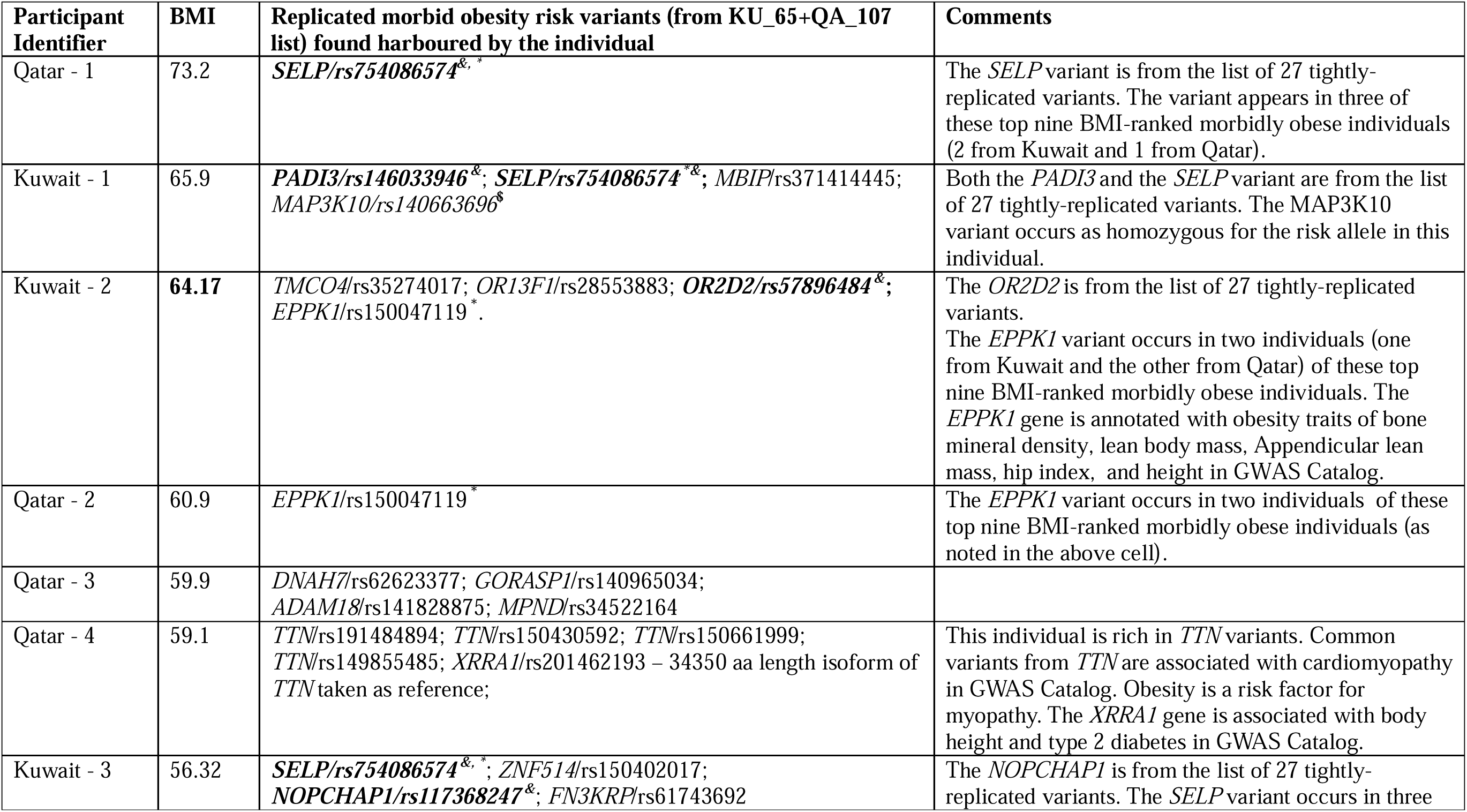
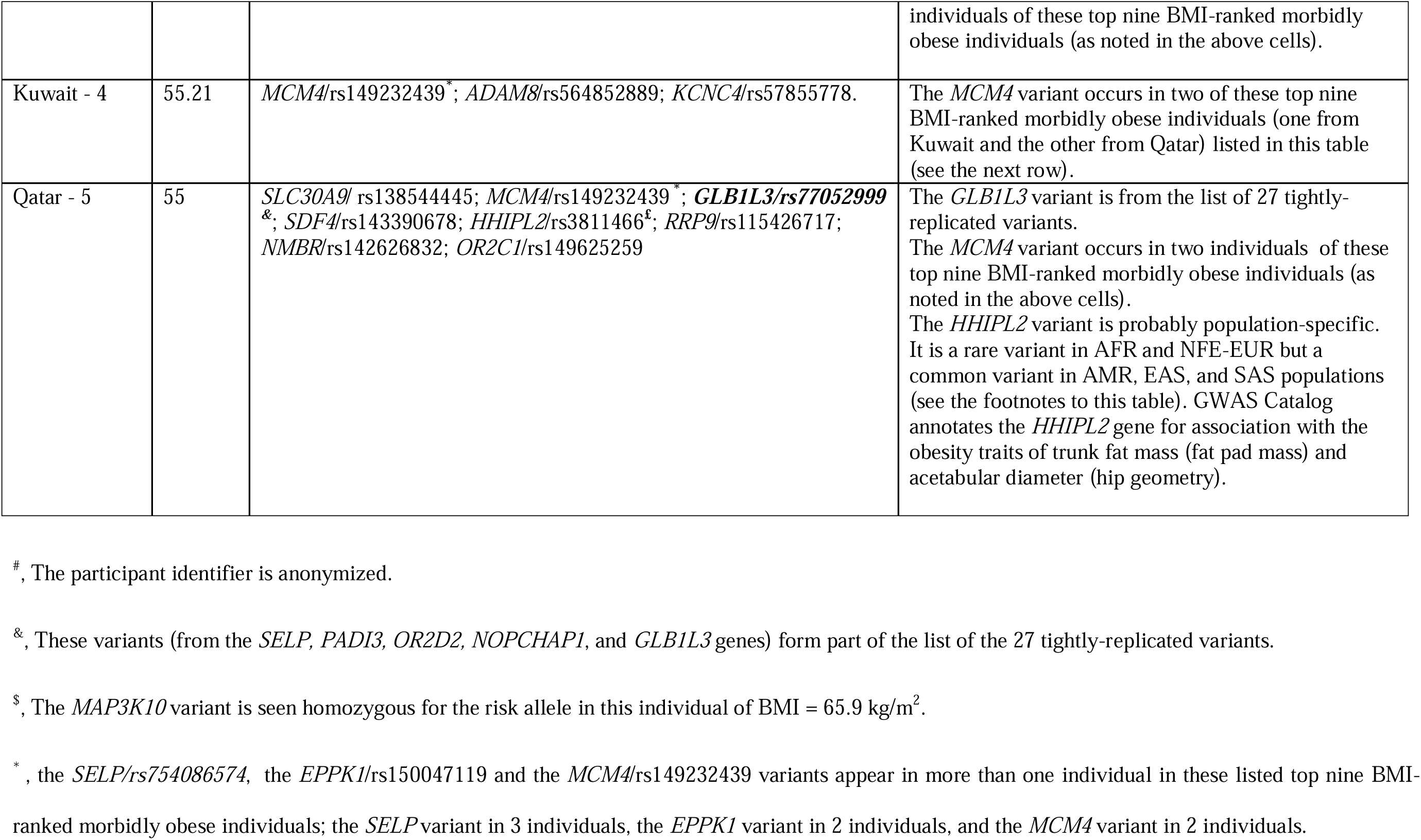
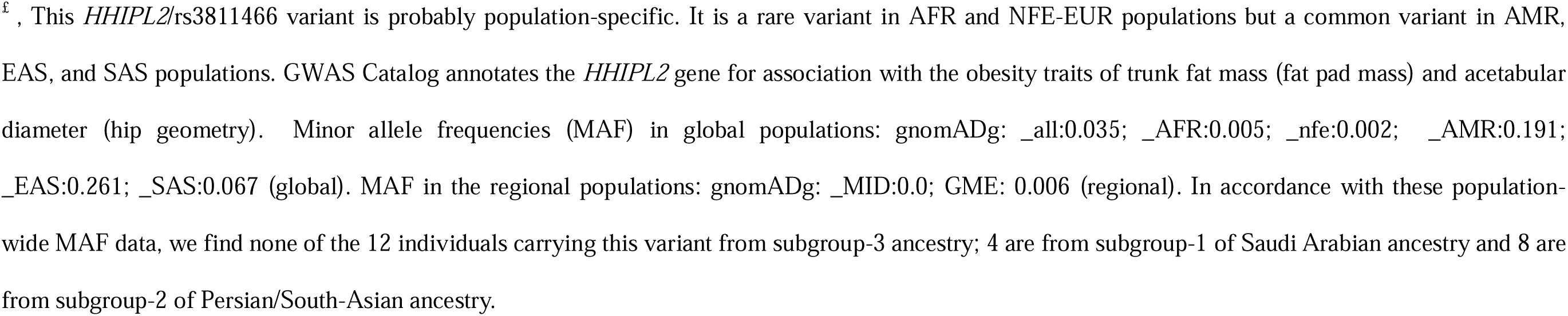
The top nine BMI-ranked morbidly obese individuals from the combined cohort and the leniently-replicated morbid obesity risk variants seen in these individuals.

### Continental population specificity of certain tightly-replicated morbid obesity variants

Examination of the MAF of the 27 tightly-replicated risk variants in regional populations (Middle East population as listed in gnomADg V4.1, and Arabian Peninsula population as listed in GME) and in global populations (as listed in gnomADg V4.1) (**Supplementary Table 3**), revealed 12 of these variants as low-frequency variants (with MAF ≥1% & ≤3.1%) and the remaining as rare variants in the region. Among the low-frequency variants, five (rs57896484/*OR2D2*, rs61746813/*ATG2A*, rs76348543/*KCNMB1*, rs77052999/*GLB1L3*, and rs61740565/*TTC32*) appeared as common variants in African population (MAF: 11%, 10.4%, 7.1%, 7%, and 5.2%, respectively); while one variant (rs78661674/*APMAP*) appeared as a common variant in East Asian population (MAF: 5.7%).

### Comparison of the identified risk gene loci with established obesity-associated genes in the GWAS Catalog^26^

As many as 201 (56%) of the 356 genes presenting the KU_374 morbid obesity candidate risk variants, 685 (64%) of the 1065 genes presenting the QA_1155 variants, 180 (63%) of the 288 genes presenting the (KU_119+QA_216) loosely-replicated risk variants, and 88 (62%) of the 142 genes presenting the (KU_65+QA_107) leniently-replicated variants were linked to obesity traits in the GWAS Catalog (Table 3). Notably, 13 of the 27 tightly-replicated gene loci were associated with direct obesity (physical/body composition) traits in the GWAS Catalog; nearly all the 27 risk gene loci were seen associated with obesity, other metabolic traits and disorders, brain morphology and brain-related disorders (Table 4).

### Homozygous variant analysis

Among the 147 leniently-replicated variants, 22 were observed as homozygous for the risk allele in one or more individuals from the combined cohort. Of these 22 variants, 8 occurred in 9 morbidly obese individuals (and 10 in 11 obese individuals, and 4 in 4 overweight individuals). Examination of the 9 morbidly obese individuals carrying these 8 homozygous variants revealed that individuals homozygous for risk allele presented generally a higher BMI compared to morbidly obese individuals carrying heterozygous genotype (**Table 6).** Specifically, the individual homozygous for risk allele at the *MAP3K10*/rs140663696 variant exhibited a BMI increase of 16 units, while individual with homozygosity for risk allele at *LRP*/rs777902155 variant showed an increase of 4 units, and the individual with homozygosity for risk allele at *ATG2A*/rs61746813 presented an increase of 2 units, compared to the highest BMI of morbidly obese individuals with heterozygosity (Table 6). Notably, the *MAP3K10* homozygous variant appeared in the individual ranked second (at a BMI of 66 kg/m^2^) among the top nine BMI-ranked morbidly obese individuals.

**Table 6.**
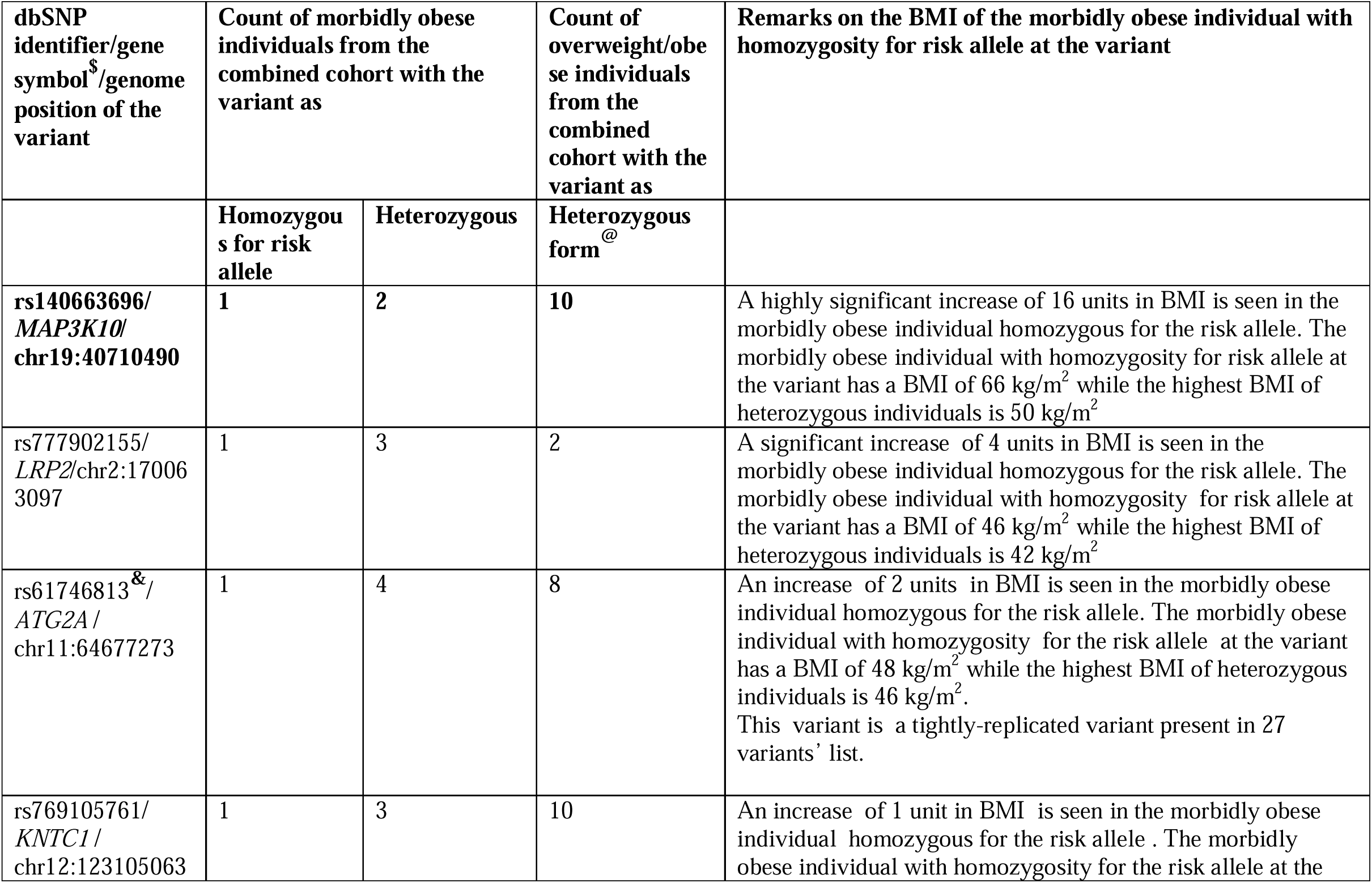
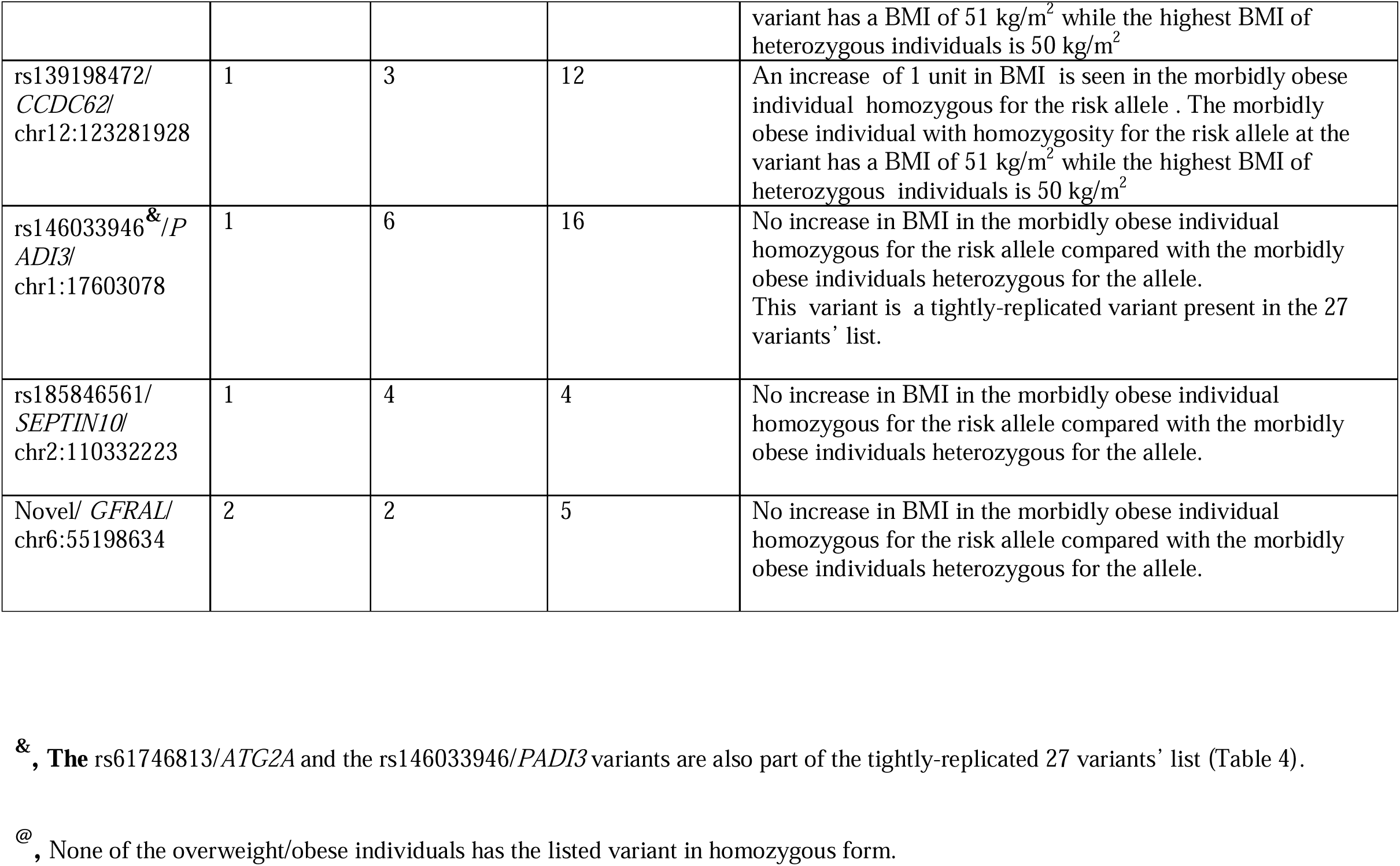
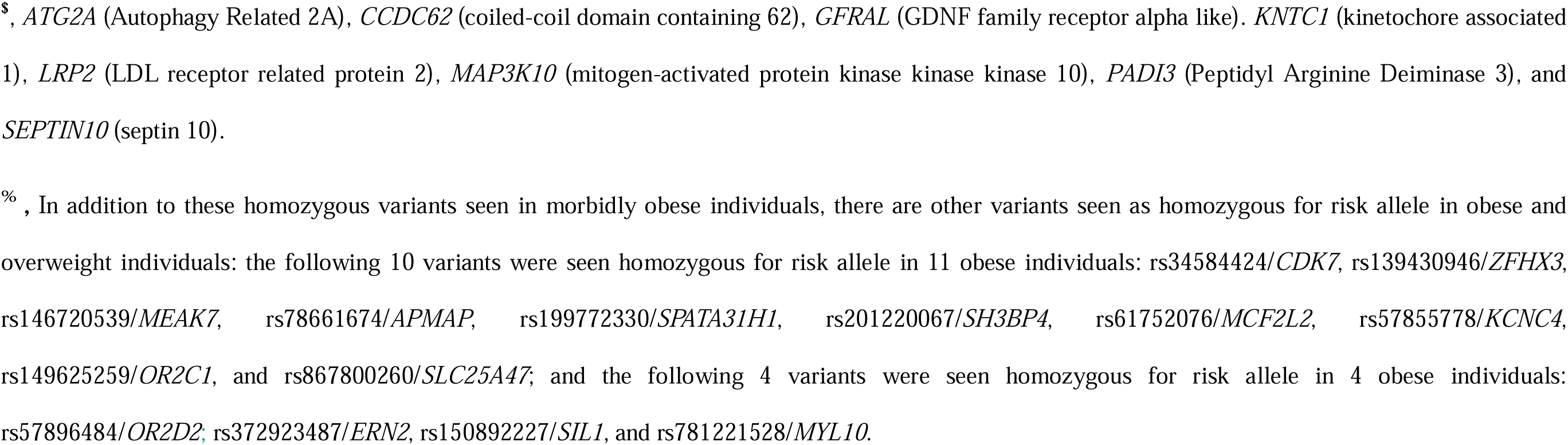
Leniently-replicated morbid obesity risk variants homozygous for risk allele^%^.

## Discussion

Polygenic obesity, a heterogeneous disorder with a spectrum of severity from overweight to obese to morbid obesity, is influenced by genetic and environmental risk factors. Identifying specific causative genetic variants has been particularly challenging for morbid obesity. In this study, we examined whole-exome sequences of >1100 Arab individuals (including 190 morbidly obese individuals) from Kuwait and Qatar to identify rare and low-frequency deleterious protein-coding variants that distinguish the morbidly obese from normal-weight individuals. The study identified an initial set of 374 and 1155 variants that preferentially occur in exomes of morbidly obese individuals from Kuwait and Qatar, respectively. These variants were further narrowed down into two categories of replication confidence: **(a)** A set of 147 Kuwait-Qatar leniently-replicated risk variants, each of which occurred in ≥2 morbidly obese individuals in one cohort and in ≥1 morbidly obese individuals in the other cohort. Up to 87 % of morbidly obese (and 46% of overweight/obese) individuals from the combined cohort exhibited at least one of these 147 variants. These variants exerted differential burden across overweight, obese and morbidly obese categories with the cumulative variant burden correlating significantly with BMI values across the obesity continuum (r=0.30, 95% CI [0.20, 0.40]; p<0.001). **(b)** A set of 27 tightly-replicated morbid obesity risk variants each of which occurred in ≥2 morbidly obese individuals in each of the two cohorts. These variants appeared in 48.94% of morbidly obese (and 27.13% of overweight/obese) individuals from the combined cohort. A significant correlation of (r=0.31, 95% CI [0.20, 0.41]; p<0.001) was observed between variant burden and BMI across the obesity continuum.

25 of the 27 genes harbouring the tightly-replicated variants were seen in GWAS Catalog as associated with obesity, metabolic disorders, brain-related disorders, brain morphology and metabolic traits (Table 4); such phenotypic features are stratified as below:: **(A) Obesity traits:** *ANTXR2, CFAP43, DNAH2, FBF1, GLB1L3, LPA, NOPCHAP1, PADI3, PIK3C2G, PLEKHG1, SLC35B2, TTC32, and ZFHX3;* **(B) Lipid traits:** *DNAH2, LPA, PLEKHG1,* and *ZFHX3;* **(C) Blood pressure traits and hypertension:** *PLEKHG1* and *WRAP73***; (D) Bone mineral density:** *APMAP, SELP,* and *PIK3C2G*; **(D) Disorders with obesity as a risk factor**: *FBF1, LPA,* and *NUP210* (Coronary artery disease), *DNAH2* and *NUP210* (Type 1 diabetes), *DNAH2, NUP210,* and *ZFHX3* (T2D), *SLC36A2* (Ischemic stroke in diabetes), and *KCNMB1* (Chronic obstructive pulmonary disease); **(E) Brain-related traits and disorders:** *MEAK7,* and *ZFHX3* (Brain morphology); *ATG2A, GLB1L3, NOPCHAP1, PIK3C2G, POLR2D,* and *SELP* (Brain-related disorders: Alzheimer’s disease, Schizophrenia, Myoneural disorders); *WRAP73,* and *CAPN5* (cognitive impairment – symptoms for developing brain-related disorders); and **(F) Olfactory receptors:** *OR13A1,* and *OR2D2*. The GWAS Catalog does not provide annotation for obesity traits in the cases of *CDK7* and *KLK7* genes; however, CDK7 is known to mediate beta-adrenergic signaling in thermogenic brown and beige fat^27^ and to regulate adipose tissue maintenance and systemic energy homeostasis^28^; and *KLK7* plays a role in adipose tissue regulation, influencing whole body energy expenditure and insulin sensitivity^29^. Of the 27 tightly-replicated variants: (i) the *LPA* variant (rs41259144) is itself annotated as associated with Lp(a) levels in GWAS Catalog – the minor allele leads to a reduction in Lp(a) levels by 14 mg/dL; It is reported that diet-induced weight loss is accompanied by an increase in Lp(a) levels in obese individuals with and without T2D^30^; and (ii) the *SLC35B2* variant (rs868023411) is itself associated with carnitine measurement – the alternate allele A is associated with lower carnitine levels; carnitine plays a substantial physiological role in lipid metabolism and intermediary metabolic pathways^31,32^.

Interestingly, the analysis identified brain-related genes and olfactory receptor genes as morbid obesity risk loci, consistent with published reports that (i) both the early-onset monogenic obesity and common polygenic obesity has a strong neurological and mental/behavioural component^12,33^: and (ii) genetic variations in olfactory receptor genes may influence the susceptibility to olfactory sensing and odors, which in turn can drive food preferences and poor eating habits, potentially leading to predisposition to obesity^34–37^.

Common variants from up to 60% of the leniently-replicated risk genes are annotated for association with obesity and anthropometric traits in GWAS Catalog. GTEx expression profiling of the identified risk genes demonstrated high transcriptional activity in adipose, skeletal muscle, pancreas, and metabolic tissues, aligning with their known roles in lipid metabolism, insulin sensitivity, and energy homeostasis. Expression profile of duodenal tissue from morbidly obese individuals and normal-weight individuals indicated 27 of the 142 leniently-replicated risk genes to exhibit significant dysregulation (p-value of ≤ 0.05) in morbidly obese individuals – particularly notable are *APMAP, MPEG1, DOCK2, SEMA4D, TTC32,* and *ZNF280D* genes that exhibited a log2FC value of ≥ 0.5 and p-value of ≤ 0.05 (Supplementary Table 3). Top pathways enriched with the identified risk genes included terms reflecting metabolic and adipogenic processes underlying obesity – white adipose tissue mass & morphology, fat cell morphology, increased total body fat, increased epididymal fat pad amount, increased body mass, decreased lean body mass, lipid metabolism, cholesterol levels, homeostasis and metabolism, insulin secretion/resistance, mTOR signaling, IL-2/STATS signaling, and p53 pathway, NAFLD, and cardiac muscle tissue morphology. Collectively, these findings underscore the functional relevance of the identified risk genes to obesity pathophysiology.

Five of the top nine BMI-ranked morbidly obese individuals harboured one or more of the tightly-replicated variants from *SELP*, *PADI3*, *OR2D2*, *NOPCHAP1*, and *GLB1L3* (Table 5); of these, the variants from *SELP*, *PADI3*, and *PADI3,* even with the limited sample sizes of the cohorts of morbidly obese individuals, exhibited statistically significant large effect sizes among the morbidly obese individuals (see Results). The SELP-rs754086574 is particularly notable as it is seen in 3 of these top nine BMI-ranked individuals with a BMI of 73.2, 65.9, and 56.4 kg/m^2^. The average BMI of the carriers of this variant among the morbidly obese individuals is 60.7 while that of non-carriers is only 44.2 kg/m^2^. The *SELP* coding for P-Selectin is involved in obesity-related chronic inflammation, and P-Selectin Glycoprotein Ligand-1 (Psgl-1), the primary leukocyte ligand for P-selectin, has been suggested to act as a cellular link between perivascular adipose inflammation and endothelial dysfunction^38^; and therapies that inhibit Psgl-1 binding to selectins have been suggested as useful to reduce obesity-associated preclinical vascular abnormalities^39^. Two other leniently-replicated variants (*EPPK1*-rs150047119 and *MCM4*-rs149232439) appear in two of these top nine BMI-ranked morbidly obese individuals (Table 5). The *EPPK1* gene is associated with obesity traits such as bone mineral density, lean body mass, appendicular lean mass, hip index, and height in the GWAS Catalog. The *MCM4* gene encodes minichromosome maintenance complex component 4 that is essential for the initiation of eukaryotic genome replication.

Further, our study highlights the relevance of homozygosity to polygenic obesity. Morbidly obese individuals homozygous for the risk allele at *MAP3K10-* rs140663696_p.T321M, and the LRP2-rs777902155_p.D2378G leniently-replicated variants present an increase in BMI (by 16 and 4 kg/m^2^, respectively) compared to morbidly obese individuals heterozygous for risk allele. Evidence from previous studies supports the relevance of homozygosity to polygenic obesity – the *FTO*-rs9939609 polymorphism has been shown to increase body weight and obesity risk in individuals carrying two copies of the risk allele^40–42^. *MAP3K10* gene is annotated in the GWAS Catalog for associations with birth weight, and glomerular filtration rate (GFR). Obesity has been associated with increased risk of GFR decline and mortality in individuals with and without chronic kidney disease^43^. Studies in mouse models of diet-induced obesity have demonstrated that *MAP3K10* (MLK2) and *MAP3K11* (MLK3) contribute to high fat diet-induced obesity by regulating energy expenditure as part of Mixed-Lineage Protein Kinase (MLK) Pathway^44^. *LRP2* gene is associated in GWAS Catalog with height, glomerular filtration rate, creatinine and cystatin C levels, and chronic kidney disease. In mouse models, LRP2, along with clusterin, has been identified as critical components of the hypothalamic feeding regulatory pathway^45^.

Both the Kuwaiti and Qatari populations are comprised of three main genetic substructures – subgroup-1 of Saudi Arabian tribe ancestry, subgroup-2 of Persian and South Asian ancestry, and subgroup-3 of nomadic tribe ancestry with a component of African ancestry; 50.4% of the individuals from the combined cohort belongs to the subgroup-1, 42.2% to subgroup-2, and 7.4% to subgroup-3. Signature of the African ancestry component is seen in the list of the identified morbid obesity risk variants – five of 27 tightly-replicated variants (namely rs57896484/*OR2D2*, rs61746813/*ATG2A*, rs76348543/*KCNMB1*, rs77052999/*GLB1L3*, and rs61740565/*TTC32*) are seen as ‘common’ variants (with MAF>5%) in the African population (**Supplementary Table 3**). Up to 39.4% of the 66 overweight/obese/morbidly obese individuals and 20% of the 20 morbidly obese individuals possessing one or more of these 5 variants are seen from African ancestry subgroup-3; these proportions are much larger than the expected proportion of 7.4.

This study has the following limitations: (i) The cohorts are of limited sample sizes; saturation in the detection of rare and low-frequency variants and performing rigorous genetic association tests would require much larger sample sets. (ii) The study cohorts are not longitudinal and hence the progression of overweight and obese individuals harbouring the risk variants to morbid obesity over time could not be assessed.

In summary, this exome-based study using Kuwaiti and Qatari cohorts identified a broad set of 147 deleterious rare and low-frequency risk variants narrowing down to a prioritised set of 27 variants with higher confidence of replication for morbid obesity. About 60% of the gene loci harbouring these variants are known to be associated with obesity and anthropometric traits, mostly BMI, in global GWA studies, and the rest of the genes with other metabolic traits, metabolic disorders, brain morphology and brain-related disorders. Analyses pertaining to pathway enrichment, tissue-specific expression enrichment, and expression data from duodenum tissues of morbidly obese individuals provided further support for these gene loci to be truly related to obesity pathogenesis These variants exhibited high penetrance in the combined cohort. Statistically significant correlations were observed between increasing variant burden and higher BMI across the continuum of polygenic obesity, from overweight to obese to morbid obesity. Based on our observations that up to 60% of the identified risk gene loci in this study have common variants annotated for association with obesity traits in the GWAS Catalog, we propose that these identified rare and low-frequency variants may act additively with GWAS BMI-associated common variants to define the total risk effect contributed by these gene loci. Such additive effects of common and rare susceptibility alleles on BMI have previously been proposed and illustrated in studies of Akbari et al. ^16^ and Zhao et al.^49^. 40% of the identified gene loci, that are not reported in global GWA studies for direct associations with obesity traits, may help to highlight new pathways contributing to obesity.

## Methods

### Ethics statement

The study was approved by the Ethical Review Committee of Dasman Diabetes institute and was carried out in accordance with the principles of the Declaration of Helsinki as revised in 2008. Written informed consent was obtained from all study participants prior to collection of blood samples. Trained nurses collected blood samples. Identities of the participants were kept confidential; samples and data were handled anonymously.

### Study cohorts: Participants recruitment and sample collection

The study included two cohorts, one from Kuwait and the other from Qatar.

The Kuwaiti study cohort comprised 285 native individuals selected from our earlier cohort of unrelated, native Kuwaiti individuals ^50^. High quality DNA samples were subjected to exome enrichment using the TruSeq Exome Enrichment kit (Illumina Inc. USA) and the Nextera Rapid Capture Exome kit (Illumina Inc. USA). The enriched libraries were subsequently clustered using the TruSeq Paired Cluster Kit V3 (Illumina Inc. USA). The sequencing process was performed on the HiSeq 2000 platform, employing Illumina’s Sequencing by Synthesis technology, resulting in 100 bp paired-end reads. Sequence reads were aligned to the human genome reference build hg19/GRCh37 using Burrows-Wheeler Aligner (BWA)^51^. Prior to variant calling, the alignment files were processed using the Genome Analysis Toolkit (GATK)^52^. Post-alignment procedures included PCR duplicate removal, local realignment around indels and base quality recalibration. Best practices for the GATK workflow were followed, and standard hard filtering parameters ^52,53^ were used for variant discovery from the processed alignment files. Variant calling was performed on BAM file from each of the samples using Haplotype Caller followed by joint genotyping analysis of the resultant gVCFs to create raw SNV and indel VCFs. To improve the quality of the data set, the resulting variant call sets were filtered by setting sample variant thresholds at 10 depth, 180 depth and genotype quality score at 20.

The study cohort from Qatar was generated by using whole-exome sequencing data of Qatari individuals publicly available from the National Center for Biotechnology Information Sequence Read Archive, with the following accessions: SRP060765, SRP061943, and SRP061463^54,55^. The Qatari individuals were sequenced using the Agilent SureSelect Human All Exon V5 and V4 kits (Agilent Technologies Inc., United States) on the Illumina HiSeq platform. We downloaded the data and considered only those exomes from native Qatari individuals (n=853) and for whom BMI values and population sub-group classifications were available from the original authors.

### Classification of the participants as normal-weight, overweight, obese and morbidly obese individuals

The study cohorts comprised 285 native individuals from Kuwait and 853 from Qatar. BMI was used to categorize these individuals as normal-weight (BMI 25 kg/m^2^) (Kuwait:n=68; Qatar:n=86), overweight (BMI 25 kg/m^2^ and < 30 kg/m^2^) (Kuwait:n=65; Qatar:n=243), obese (BMI 30 kg/m^2^ and < 40 kg/m^2^) (Kuwait:n=85; Qatar:n=403) and morbidly obese (BMI 40 kg/m^2^) (Kuwait:n=67; Qatar:n=123).

### Deriving the morbid obesity specific variants

In accordance with the methodology outlined in **Supplementary Fig. 1** and using Annovar^56^, a systematic variant filtering process was executed. Variants were first filtered by a minimum read depth (DP) of 8X and a mapping quality (MQ) ≥ 40. High-quality variants were further restricted to autosomal single nucleotide polymorphisms (SNPs) observed in at least two morbidly obese individuals and absent in normal-weight controls. Furthermore, only variants with missense or loss-of-function (LOF) consequences were retained. To enhance the specificity of our analysis, a dual population frequency filter was employed. Initially, variants with a population minor allele frequency (MAF) exceeding 0.05 in the Exome Aggregation Consortium (ExAC) database^57^ (http://exac.broadinstitute.org/) were excluded. Subsequently, a more refined filter was applied, retaining variants with an MAF 0.05 in the Arabian Peninsula (AP) subset of the Greater Middle Eastern (GME) Variome database^22^. In the ensuing step, a stringent functionality filter was implemented to exclude variants characterized as non-damaging (‘tolerated’ or ‘benign’) by SIFT ^58^ and PolyPhen ^59^ tools. As an additional step, the resulting variants were filtered based on their Combined Annotation-Dependent Depletion score ^60^ to prioritise functional, and deleterious variants across many functional categories, effect sizes and genetic architectures. A scaled score of ≥ 20 was applied to retain only those variants that were predicted to be among the top 1% of deleterious variants in the human genome. The variants were also annotated for pathogenic significance using OMIM^61^ and ClinVar^62^ As is the practice, in cases of variants with conflicting annotation for clinical significance in ClinVar, evidence from peer-reviewed publications and from OMIM took precedence over evidence from clinical testing submissions^63^.

### Validating the morbid obesity risk variants for replication between the two study cohorts

Replication of morbid obesity-associated variants between the two study cohorts was assessed using three approaches as illustrated in **Supplementary Fig. 3**. **Approach 1 – Loosely-replicated variants**: A variant identified in morbidly obese individuals from one cohort is considered loosely-replicated if it is present in at least one morbidly obese or overweight/obese individual and absent in all normal-weight individuals in the other cohort. **Approach 2 – Leniently-replicated variants**: A morbid obesity-associated variant identified from one cohort is defined as leniently-replicated, if it occurs in at least one morbidly obese individual from the other cohort and is absent in normal-weight individuals in the other cohort. **Approach 3 – tightly-replicated variants**: A variant is classified as tightly-replicated if it is seen in two or more morbidly obese individuals (and absent in normal-weight individuals) in each of the two cohorts from Kuwait and Qatar.

### Genetic association tests on the tightly-replicated variants with BMI

BMI association analysis was performed in the extreme phenotype subset comprising normal-weight and morbidly obese individuals, with BMI modelled as a continuous trait. Associations with a p-value < 0.05 were considered as significant.

### Variant burden and BMI correlation analysis

Correlations between the body mass index and count of morbid obesity risk variants were examined using Pearson’s product-moment correlation tests. We considered a threshold of <0.001 for p-value to consider the correlation as statistically significant. The models for correlations were adjusted for age, sex, and the population genetic substructure stratifications.

### Examining the identified risk gene loci against the NHGRI-EBI GWAS Catalog

We used the NHGRI-EBI GWAS Catalog^26^ to examine the gene loci, that we identify in the study, for known associations with obesity traits through common variants in global studies. For this purpose, we queried the GWAS Catalog (as on 6 August 2025) using a multitude of obesity-related keywords – such as body mass index, obesity, waist circumference, waist-to-hip ratio, height, weight, adiposity, fat mass, fat, and percentage body fat.

### Pathway and tissue enrichment analysis

To delineate the underlying biological processes and pathways impacted by the identified gene loci, we performed pathway and tissue enrichment analysis. We performed biological process enrichment analysis by using the DAVID^64^ and Enrichr ^65^ tools. In addition, we used the KEGG^66^, Reactome ^67^ and wiki pathway ^68^ databases to perform pathway enrichment analysis of the gene loci identified in morbidly obese participants. The most statistically enriched gene ontology (GO) terms were visualized using the ggplot2 visualization package ^69^. Furthermore, we performed gene-based tissue expression enrichment for genes, harbouring the identified variants, in adult tissues from morbidly obese individuals, by utilizing data from the Genotype-Tissue Expression (GTEx) database (www.gtexportal.org) [portal last accessed on Feb 19, 2024]. Expression data used were gene-level transcripts per million (TPM) quantifications generated by the GTEx consortium ^24^.

### Acquisition of publicly available expression profiles of duodenum from morbidly obese individuals

Ho-Plágaro et al.^25^ generated transcription profiling of duodenum from morbidly obese individuals and non-obese individuals from Spain with an aim to identify intestinal genes that may be associated with the development of morbid obesity in women regardless of the degree of insulin resistance of patients. The dataset is publicly available in the Gene Expression Omnibus (GEO) resource with the accession identifier of GSE147562. Raw count data were downloaded and processed using the *DESeq2* pipeline in R package. Samples were stratified into non-obese, and morbidly obese groups, with additional subdivision based on insulin resistance status. Quality control, normalization, and variance-stabilizing transformation were applied prior to performing differential expression analysis. Differentially expressed genes (DEGs) between groups were identified using a Wald test with Benjamini–Hochberg correction, and genes with an adjusted p-value < 0.05 and |log2FC| ≥ 0.5 were considered significant. Bar plots were generated using ggplot2^69^ and pheatmap^70^ to visualize expression patterns of the 27 tightly-replicated obesity-associated genes.

## Data Availability

The generated datasets of morbid obesity variants are presented as Supplementary Datasets D1-D4. Dataset D1 corresponds to the KU_374 variants list; D2 corresponds to the QA_1155 variants list; Dataset D3 corresponds to the KU_119+QA_216 loosely-replicated variants list; Dataset D4 corresponds to the KU_65+QA_107 leniently-replicated variants list.

## Acknowledgements

Members of the BioBank Facility at Dasman Diabetes Institute are acknowledged for sample processing and DNA extraction.

## Author Contributions

T.A.T. performed the study design for this work and directed the study components. S.E.J. processed all the sequenced exomes. S.E.J., A.C. and Md.Z.M performed all the data analysis and participated in interpreting the results. T.A.T. and S.E.J. developed the manuscript draft. A.C. and Md.Z.M. contributed to developing the manuscript. T.A.T. reviewed and finalized the manuscript. A.A.M. critically reviewed the manuscript and participated in discussions. F.A-M. provided the required resources, participated in discussions, critically reviewed and approved the manuscript.

## Competed Interests

The authors declare no competing interests.

## References

1 GBD 2021 Adult BMI Collaborators. Global, regional, and national prevalence of adult overweight and obesity, 1990-2021, with forecasts to 2050: a forecasting study for the Global Burden of Disease Study 2021. Lancet 405, 813–838, doi:10.1016/S0140-6736(25)00355-1 (2025).

2 GBD 2021 Adult BMI Collaborators. Global, regional, and national prevalence of child and adolescent overweight and obesity, 1990-2021, with forecasts to 2050: a forecasting study for the Global Burden of Disease Study 2021. Lancet 405, 785–812, doi:10.1016/S0140-6736(25)00397-6 (2025).

3 Okunogbe, A. et al. Economic impacts of overweight and obesity: current and future estimates for 161 countries. BMJ Glob Health 7, doi:10.1136/bmjgh-2022-009773 (2022).

4 Al-Sabah, S. et al. The economic impact of obesity in Kuwait: a micro-costing study evaluating the burden of obesity-related comorbidities. J Med Econ 26, 1368–1376, doi:10.1080/13696998.2023.2265721 (2023).

5 Butalia, S. et al. Health care cost of severe obesity and obesity-related comorbidities: A retrospective cohort study from Alberta, Canada. Obes Res Clin Pract 17, 421–427, doi:10.1016/j.orcp.2023.09.006 (2023).

6 Emmerich, S. D., Fryar, C. D., Stierman, B. & Ogden, C. L. Obesity and Severe Obesity Prevalence in Adults: United States, August 2021-August 2023. NCHS Data Brief, doi:10.15620/cdc/159281 (2024).

7 Channanath, A. M., Farran, B., Behbehani, K. & Thanaraj, T. A. State of diabetes, hypertension, and comorbidity in Kuwait: showcasing the trends as seen in native versus expatriate populations. Diabetes care 36, e75, doi:10.2337/dc12-2451 (2013).

8 Bouchard, C. Genetics of Obesity: What We Have Learned Over Decades of Research. Obesity 29, 802–820, doi:10.1002/oby.23116 (2021).

9 Kunzel, R. et al. Detecting monogenic obesity: a systematic exome-wide workup of over 500 individuals. Int J Obes (Lond) 49, 1400–1411, doi:10.1038/s41366-025-01819-0 (2025).

10 Khera, A. V. et al. Polygenic Prediction of Weight and Obesity Trajectories from Birth to Adulthood. Cell 177, 587–596 e589, doi:10.1016/j.cell.2019.03.028 (2019).

11 Kavaja, F., Liehr, T. & Temaj, G. Genetic architecture of obesity and advances in precision pharmacotherapy: a comprehensive review. Acta Biochim Pol 73, 15484, doi:10.3389/abp.2026.15484 (2026).

12 Loos, R. J. F. & Yeo, G. S. H. The genetics of obesity: from discovery to biology. Nat Rev Genet 23, 120–133, doi:10.1038/s41576-021-00414-z (2022).

13 Smit, R. A. J. et al. Polygenic prediction of body mass index and obesity through the life course and across ancestries. Nat Med 31, 3151–3168, doi:10.1038/s41591-025-03827-z (2025).

14 Turcot, V. et al. Protein-altering variants associated with body mass index implicate pathways that control energy intake and expenditure in obesity. Nature genetics 50, 26–41, doi:10.1038/s41588-017-0011-x (2018).

15 Antoine, D. et al. Low-frequency Coding Variants Associated With Body Mass Index Affect the Success of Bariatric Surgery. The Journal of clinical endocrinology and metabolism 107, e1074–e1084, doi:10.1210/clinem/dgab774 (2022).

16 Akbari, P. et al. Sequencing of 640,000 exomes identifies GPR75 variants associated with protection from obesity. Science 373, doi:10.1126/science.abf8683 (2021).

17 Jiao, H. et al. Exome sequencing followed by genotyping suggests SYPL2 as a susceptibility gene for morbid obesity. European journal of human genetics: EJHG 23, 1216–1222, doi:10.1038/ejhg.2014.255 (2015).

18 Jiao, H. et al. Whole-Exome Sequencing Suggests LAMB3 as a Susceptibility Gene for Morbid Obesity. Diabetes 65, 2980–2989, doi:10.2337/db16-0522 (2016).

19 Saeed, S., Arslan, M. & Froguel, P. Genetics of Obesity in Consanguineous Populations: Toward Precision Medicine and the Discovery of Novel Obesity Genes. Obesity 26, 474–484, doi:10.1002/oby.22064 (2018).

20 Hebbar, P. et al. Genome-wide association study identifies novel recessive genetic variants for high TGs in an Arab population. J Lipid Res 59, 1951–1966, doi:10.1194/jlr.P080218 (2018).

21 Younes, S., Ibrahim, A., Al-Jurf, R. & Zayed, H. Genetic polymorphisms associated with obesity in the Arab world: a systematic review. Int J Obes (Lond) 45, 1899–1913, doi:10.1038/s41366-021-00867-6 (2021).

22 Scott, E. M. et al. Characterization of Greater Middle Eastern genetic variation for enhanced disease gene discovery. Nature genetics 48, 1071–1076, doi:10.1038/ng.3592 (2016).

23 Chen, S. et al. A genomic mutational constraint map using variation in 76,156 human genomes. Nature 625, 92–100, doi:10.1038/s41586-023-06045-0 (2024).

24 Lonsdale, J. et al. The genotype-tissue expression (GTEx) project. Nature genetics 45, 580–585 (2013).

25 Ho-Plagaro, A. et al. Morbid Obesity in Women Is Associated with an Altered Intestinal Expression of Genes Related to Cancer Risk and Immune, Defensive, and Antimicrobial Response. Biomedicines 10, doi:10.3390/biomedicines10051024 (2022).

26 Cerezo, M. et al. The NHGRI-EBI GWAS Catalog: standards for reusability, sustainability and diversity. Nucleic acids research 53, D998–D1005, doi:10.1093/nar/gkae1070 (2025).

27 Ji, H. et al. CDK7 Mediates the Beta-Adrenergic Signaling in Thermogenic Brown and White Adipose Tissues. iScience 23, 101163, doi:10.1016/j.isci.2020.101163 (2020).

28 Chen, Y. et al. Adipocyte-specific CDK7 ablation leads to progressive loss of adipose tissue and metabolic dysfunction. FEBS letters 596, 1434–1444, doi:10.1002/1873-3468.14335 (2022).

29 Zieger, K. et al. Ablation of kallikrein 7 (KLK7) in adipose tissue ameliorates metabolic consequences of high fat diet-induced obesity by counteracting adipose tissue inflammation in vivo. Cell Mol Life Sci 75, 727–742, doi:10.1007/s00018-017-2658-y (2018).

30 Berk, K. A. et al. Effect of diet-induced weight loss on lipoprotein(a) levels in obese individuals with and without type 2 diabetes. Diabetologia 60, 989–997, doi:10.1007/s00125-017-4246-y (2017).

31 Adeva-Andany, M. M., Calvo-Castro, I., Fernandez-Fernandez, C., Donapetry-Garcia, C. & Pedre-Pineiro, A. M. Significance of l-carnitine for human health. IUBMB Life 69, 578–594, doi:10.1002/iub.1646 (2017).

32. Dahash, B. A. & Sankararaman, S. in StatPearls Ch. Carnitine Deficiency, (2025).

33 Saeed, S., Bonnefond, A. & Froguel, P. Obesity: exploring its connection to brain function through genetic and genomic perspectives. Mol Psychiatry 30, 651–658, doi:10.1038/s41380-024-02737-9 (2025).

34 Jarick, I. et al. Novel common copy number variation for early onset extreme obesity on chromosome 11q11 identified by a genome-wide analysis. Human molecular genetics 20, 840–852, doi:10.1093/hmg/ddq518 (2011).

35 Choquette, A. C. et al. Association between olfactory receptor genes, eating behavior traits and adiposity: results from the Quebec Family Study. Physiol Behav 105, 772–776, doi:10.1016/j.physbeh.2011.10.015 (2012).

36 Yeomans, M. R. Olfactory influences on appetite and satiety in humans. Physiol Behav 87, 800–804, doi:10.1016/j.physbeh.2006.01.029 (2006).

37 Spinelli, S. & Monteleone, E. Food Preferences and Obesity. Endocrinol Metab (Seoul) 36, 209–219, doi:10.3803/EnM.2021.105 (2021).

38 Lee, W. J., Chait, A. & Kim, F. P-selectin glycoprotein ligand-1: a cellular link between perivascular adipose inflammation and endothelial dysfunction. Diabetes 61, 3070–3071, doi:10.2337/db12-1036 (2012).

39 Wang, H. et al. Obesity-induced endothelial dysfunction is prevented by deficiency of P-selectin glycoprotein ligand-1. Diabetes 61, 3219–3227, doi:10.2337/db12-0162 (2012).

40 Frayling, T. M. et al. A common variant in the FTO gene is associated with body mass index and predisposes to childhood and adult obesity. Science 316, 889–894, doi:10.1126/science.1141634 (2007).

41 Bressan Rente Ferreira Marum, A. et al. Association of homozygous fat mass and obesity-associated (FTO rs9939609) gene with body mass, body mass index (BMI), and the binge eating scale in women: A cross-sectional study. Clin Nutr ESPEN 69, 225–232, doi:10.1016/j.clnesp.2025.07.016 (2025).

42 Church, C. et al. Overexpression of Fto leads to increased food intake and results in obesity. Nature genetics 42, 1086–1092, doi:10.1038/ng.713 (2010).

43 Chang, A. R. et al. Adiposity and risk of decline in glomerular filtration rate: meta-analysis of individual participant data in a global consortium. BMJ 364, k5301, doi:10.1136/bmj.k5301 (2019).

44 Kant, S. et al. Role of the mixed-lineage protein kinase pathway in the metabolic stress response to obesity. Cell Rep 4, 681–688, doi:10.1016/j.celrep.2013.07.019 (2013).

45 Gil, S. Y. et al. Clusterin and LRP2 are critical components of the hypothalamic feeding regulatory pathway. Nature communications 4, 1862, doi:10.1038/ncomms2896 (2013).

46 Alsmadi, O. et al. Genetic substructure of Kuwaiti population reveals migration history. PloS one 8, e74913, doi:10.1371/journal.pone.0074913 (2013).

47 Eaaswarkhanth, M. et al. Unraveling a fine-scale high genetic heterogeneity and recent continental connections of an Arabian Peninsula population. European journal of human genetics: EJHG 30, 307–319, doi:10.1038/s41431-021-00861-6 (2022).

48 Hunter-Zinck, H. et al. Population genetic structure of the people of Qatar. Am J Hum Genet 87, 17–25, doi:10.1016/j.ajhg.2010.05.018 (2010).

49 Zhao, Y. et al. Protein-truncating variants in BSN are associated with severe adult-onset obesity, type 2 diabetes and fatty liver disease. Nature genetics 56, 579–584, doi:10.1038/s41588-024-01694-x (2024).

50 John, S. E. et al. Assessment of coding region variants in Kuwaiti population: implications for medical genetics and population genomics. Scientific reports 8, 16583 (2018).

51 Li, H. & Durbin, R. Fast and accurate short read alignment with Burrows-Wheeler transform. Bioinformatics 25, 1754–1760, doi:10.1093/bioinformatics/btp324 (2009).

52 DePristo, M. A. et al. A framework for variation discovery and genotyping using next-generation DNA sequencing data. Nature genetics 43, 491–498 (2011).

53 der Auwera, V. From FastQ data to high confidence variant calls: the Genome Analysis Toolkit best practices pipeline. Curr Protoc Bioinformatics 43, 1 (2013).

54 Rodriguez-Flores, J. L. et al. Indigenous Arabs are descendants of the earliest split from ancient Eurasian populations. Genome Res 26, 151–162, doi:10.1101/gr.191478.115 (2016).

55 Fakhro, K. A. et al. The Qatar genome: a population-specific tool for precision medicine in the Middle East. Hum Genome Var 3, 16016, doi:10.1038/hgv.2016.16 (2016).

56 Wang, K., Li, M. & Hakonarson, H. ANNOVAR: functional annotation of genetic variants from high-throughput sequencing data. Nucleic acids research 38, e164, doi:10.1093/nar/gkq603 (2010).

57 Lek, M. et al. Analysis of protein-coding genetic variation in 60,706 humans. Nature 536, 285–291, doi:10.1038/nature19057 (2016).

58 Kumar, P., Henikoff, S. & Ng, P. C. Predicting the effects of coding non-synonymous variants on protein function using the SIFT algorithm. Nature protocols 4, 1073–1081 (2009).

59 Adzhubei, I. A. et al. A method and server for predicting damaging missense mutations. Nature methods 7, 248–249 (2010).

60 Kircher, M. et al. A general framework for estimating the relative pathogenicity of human genetic variants. Nature genetics 46, 310–315 (2014).

61 Amberger, J., Bocchini, C. A., Scott, A. F. & Hamosh, A. McKusick’s Online Mendelian Inheritance in Man (OMIM). Nucleic acids research 37, D793–796, doi:10.1093/nar/gkn665 (2009).

62 Landrum, M. J. et al. ClinVar: public archive of interpretations of clinically relevant variants. Nucleic acids research 44, D862–D868 (2016).

63 Tarailo-Graovac, M., Zhu, J. Y. A., Matthews, A., Van Karnebeek, C. D. & Wasserman, W. W. Assessment of the ExAC data set for the presence of individuals with pathogenic genotypes implicated in severe Mendelian pediatric disorders. Genetics in medicine 19, 1300–1308 (2017).

64 Sherman, B. T. et al. DAVID: a web server for functional enrichment analysis and functional annotation of gene lists (2021 update). Nucleic acids research 50, W216–W221, doi:10.1093/nar/gkac194 (2022).

65 Kuleshov, M. V. et al. Enrichr: a comprehensive gene set enrichment analysis web server 2016 update. Nucleic acids research 44, W90–W97 (2016).

66 Kanehisa, M., Furumichi, M., Sato, Y., Matsuura, Y. & Ishiguro-Watanabe, M. KEGG: biological systems database as a model of the real world. Nucleic acids research 53, D672-D677, doi:10.1093/nar/gkae909 (2025).

67 Fabregat, A. et al. The reactome pathway knowledgebase. Nucleic acids research 46, D649–D655 (2018).

68 Kutmon, M. et al. WikiPathways: capturing the full diversity of pathway knowledge. Nucleic acids research 44, D488–D494 (2016).

69 Wickham, H. ggplot2. WIREs Computational Statistics 3, 180–185, 10.1002/wics.147 (2011).

70 pheatmap: Pretty Heatmaps v. 1.0.13 (GitHub, 2025).

