## Supplementary Figures Tables for "Genetic Variant Burden Rises with BMI: Insights from Rare and Low-Frequency Variants in Obesity"

**Supplementary Fig. 1. Pipeline for the derivation of morbid obesity risk variants.**

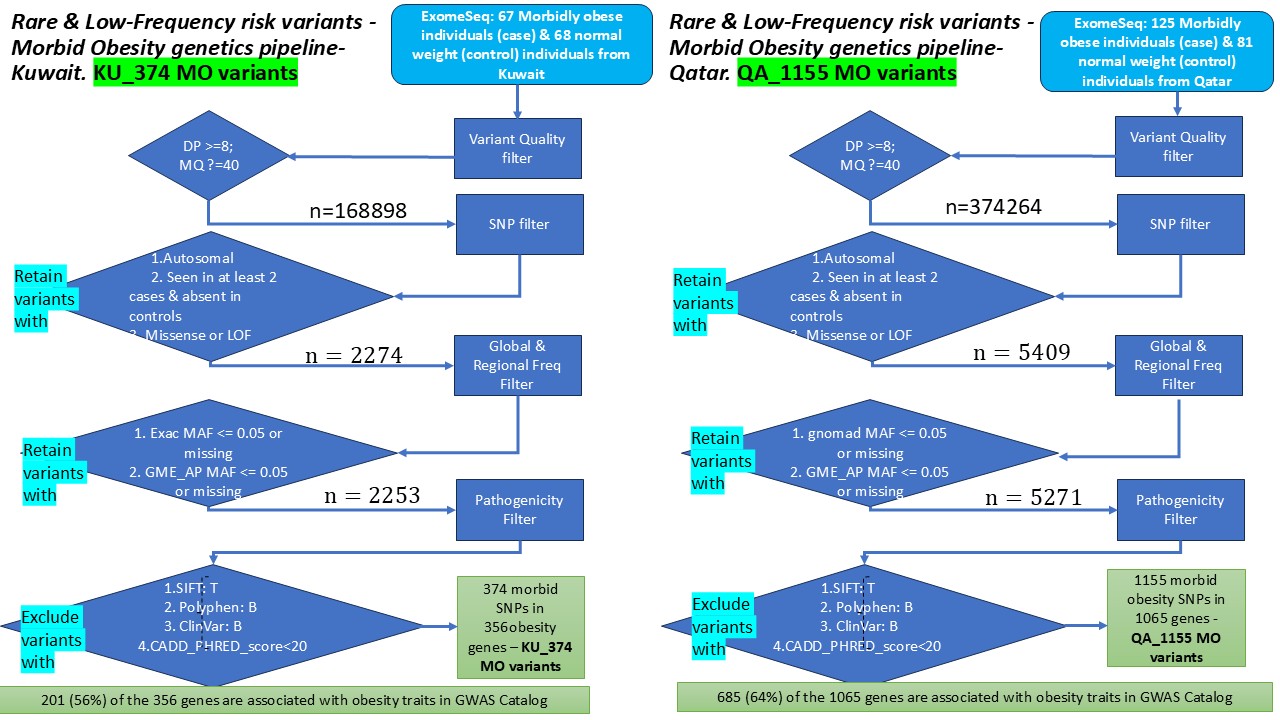

**Supplementary Fig. 2. Distribution of the allele frequencies of the morbid obesity risk variants from the KU_374 and QA_1155 variants’ lists – as seen in GME_AP (Arabian peninsula) and in gnomADg v.2.1.1 databases.**

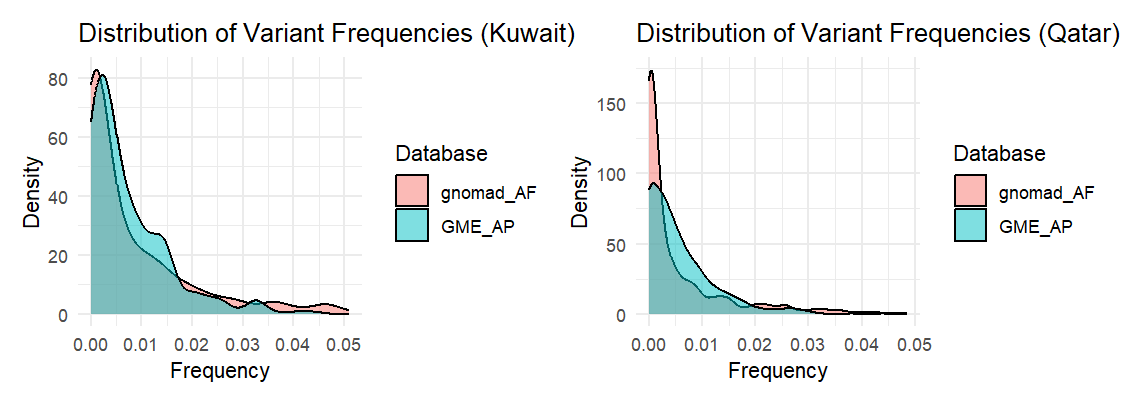

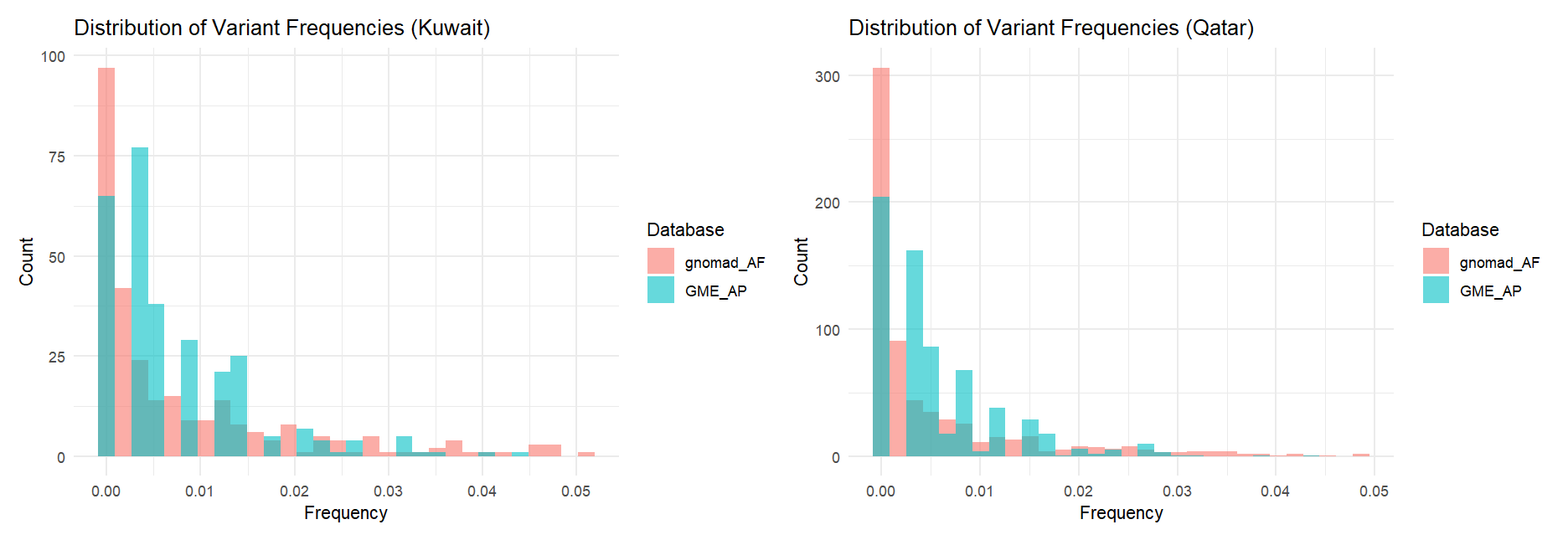

**Supplementary Fig. 3. Pipeline for validating the identified morbid obesity risk variants by checking for replication between the Kuwaiti and Qatari cohorts.**

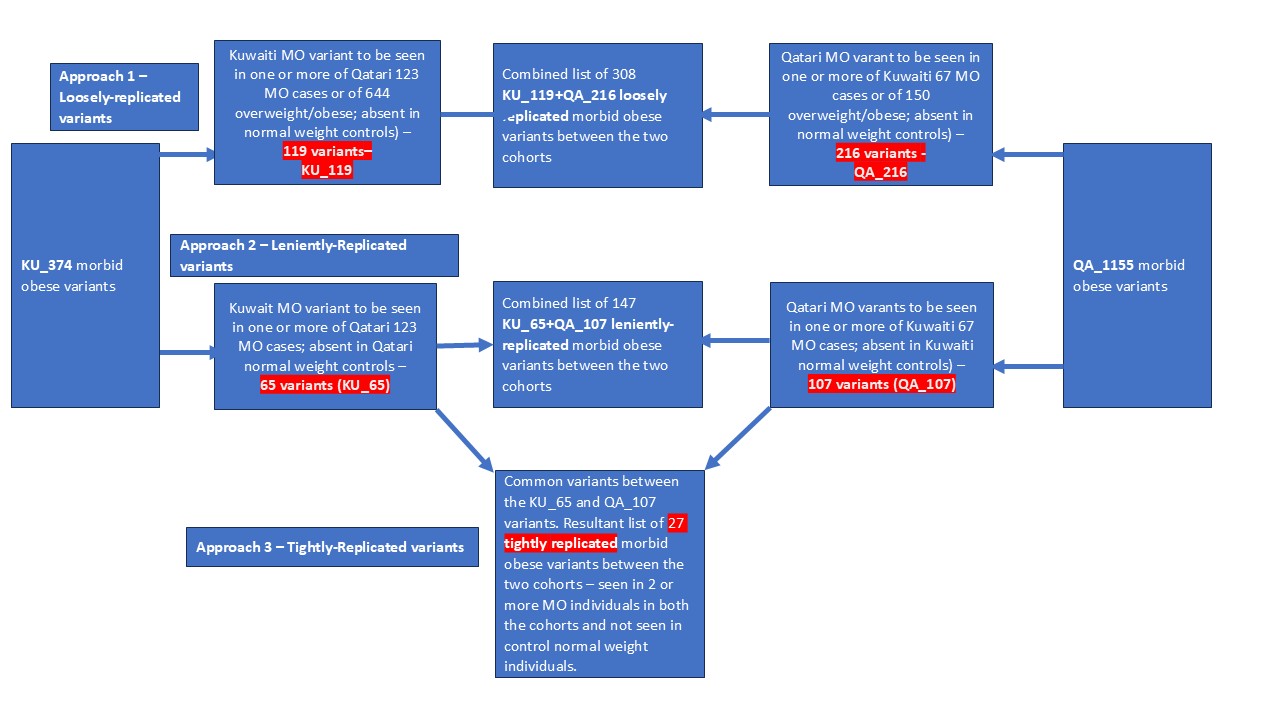

**Supplementary Table 1. Expression analysis using the publicly available GEO (GSE147562) of the 27 tightly-replicated morbid obesity risk gene loci in duodenal tissue from morbidly obese individuals compared to non-obese individuals.**

| **GENE** | **P.Value** | **LogFC** |
| --- | --- | --- |
| APMAP | 0.0060509 | 0.50773844 |
| CDK7 | 0.0096256 | -0.33066864 |
| NUP210 | 0.02825 | 0.42035858 |
| PLEKHG1 | 0.0335782 | 0.48824254 |
| PIK3C2G | 0.0367246 | -0.23097653 |
| TTC32 | 0.0412469 | -0.60624727 |

**Supplementary Table 2. Expression analysis using the publicly available GEO (GSE147562) of the 142 leniently-replicated morbid obesity risk gene loci (KU_65+QA_107 variants’ list) in duodenal tissue from morbidly obese individuals compared to non-obese individuals.**

| **Gene** | **P.Value** | **logFC** |
| --- | --- | --- |
| HIVEP3 | 0.003063 | 0.39925267 |
| APMAP | 0.006051 | 0.50773844 |
| CDK5RAP1 | 0.006362 | 0.4114596 |
| DOCK2 | 0.007944 | 0.57286372 |
| ZNF280D | 0.008739 | -0.51233354 |
| SEMA4D | 0.009219 | 0.55531569 |
| CDK7 | 0.009626 | -0.33066864 |
| MAP3K10 | 0.010119 | -0.41582365 |
| MYCBPAP | 0.014835 | -0.33775528 |
| CYP4A22 | 0.01631 | 0.35384534 |
| FN3KRP | 0.018034 | 0.48956779 |
| ZCCHC2 | 0.0202 | 0.39468274 |
| BFSP1 | 0.025264 | -0.45125953 |
| MAP1A | 0.025372 | 0.30789881 |
| NUP210 | 0.02825 | 0.42035858 |
| PEAK1 | 0.030991 | -0.3485943 |
| SLC30A9 | 0.031219 | 0.32075167 |
| KRT8 | 0.031222 | 0.44713376 |
| FAM221A | 0.031507 | -0.43164477 |
| PLEKHG1 | 0.033578 | 0.48824254 |
| CCDC40 | 0.035223 | -0.37831484 |
| PIK3C2G | 0.03672 | -0.2309765 |
| TTC32 | 0.041247 | -0.60624727 |
| METTL17 | 0.044543 | -0.2764811 |
| MPEG1 | 0.046597 | 0.74845755 |
| SH3BP4 | 0.048645 | 0.28498193 |
| CLSTN2 | 0.050839 | 0.48621247 |

**Supplementary Table 3. Global and regional allele frequencies of the 27 tightly-replicated morbid obesity risk variants.**

|  | **Global - gnomADg v4.1** | | | | | | **Regional** | |
| --- | --- | --- | --- | --- | --- | --- | --- | --- |
| **dbSNP identifier/gene symbol** | **ALL^&,@^** | **AFR^%^** | **AMR** | **EAS^%^** | **NFE=EUR** | **SAS** | **gnomADg v4.1_MID^&,$^** | **GME_AP^&,$^** |
| rs185151361/*ANTXR2* | 0.0004 | 0.0001 | 0.001 | 0 | 0.0005 | 0.0004 | 0 | 0 |
| rs78661674/*APMAP* | ***0.02*** | 0.007 | 0.019 | *0.057* | 0.024 | 0.029 | 0 | ***0.0233*** |
| rs61746813/*ATG2A* | ***0.03*** | *0.104* | 0.013 | 0.0002 | 0.001 | 0 | 0.007 | ***0.0146*** |
| rs201701148/*CAPN5* | 0.0003 | 0.0007 | 0 | 0 | 0 | 0.0002 | 0 | ***0.009*** |
| rs34584424/*CDK7* | ***0.016*** | 0.004 | 0.013 | 0.0002 | 0.025 | 0.004 | ***0.017*** | ***0.0117*** |
| rs148204337/*CFAP43* | 0.002 | 0.001 | 0.001 | 0.0002 | 0.003 | 0.002 | ***0.017*** | 0.0029 |
| rs141149043/DNAH2 | 0.003 | 0.0003 | 0.001 | 0.0001 | 0.001 | 0.019 | 0 | 0 |
| rs113062332/FBF1 | ***0.019*** | 0.005 | 0.01 | 0.0002 | 0.031 | 0.006 | 0 | 0 |
| rs77052999/**GLB1L3** | ***0.02*** | *0.07* | 0.008 | 0 | 0.0003 | 0 | ***0.014*** | ***0.009*** |
| rs76348543/KCNMB1 | ***0.02*** | *0.071* | 0.007 | 0 | 0.0003 | 0.0002 | 0.003 | 0 |
| rs112145183/KLK14 | 0.007 | 0.017 | 0.005 | 0 | 0.004 | 0.003 | ***0.014*** | 0.003 |
| rs41259144/LPA | 0.007 | 0.002 | 0.017 | 0.0002 | 0.009 | 0.0002 | 0.007 | 0.006 |
| rs146720539/MEAK7 | 0.0002 | 0.0001 | 0.0002 | 0 | 0.0002 | 0.0004 | 0.007 | 0.006 |
| rs117368247/NOPCHAP1 | ***0.019*** | 0.005 | 0.013 | 0.0002 | 0.027 | 0.005 | ***0.017*** | 0.006 |
| rs151008831/NUP210 | 0.003 | 0.001 | 0.001 | 0 | 0.005 | 0.004 | 0.003 | ***0.012*** |
| rs116719900/OR13A1 | 0.004 | 0.015 | 0.002 | 0 | 0.00004 | 0 | 0 | 0.001 |
| rs57896484/**OR2D2** | ***0.031*** | *0.11* | 0.009 | 0 | 0.0004 | 0.0004 | 0 | ***0.012*** |
| rs146033946/PADI3 | 0.0002 | 0 | 0.00006 | 0 | 0.00004 | 0 | 0 | 0.003 |
| rs146312199/PIK3C2G | 0.004 | 0.001 | 0.005 | 0 | 0.004 | 0.004 | 0 | 0.006 |
| rs117898584/PLEKHG1 | 0.001 | 0.002 | 0.001 | 0 | 0.0004 | 0 | ***0.01*** | 0.006 |
| rs142734461/POLR2D | 0.00012 | 0.00005 | 0.00032 | 0 | 0.00006 | 0 | 0.007 | ***0.009*** |
| rs754086574/SELP | 0.00007 | 0 | 0.0004 | 0 | 0.00007 | 0 | 0.003 | 0.003 |
| rs868023411/SLC35B2 | 0.00002 | 0 | 0.00007 | 0 | 0 | 0 | 0.007 | ***0.015*** |
| rs77010315/SLC36A2 | 0.009 | 0.002 | 0.007 | 0 | 0.013 | 0.003 | 0.007 | 0 |
| rs61740565/TTC32 | ***0.015*** | *0.052* | 0.004 | 0 | 0.0003 | 0.0002 | 0.007 | 0.003 |
| rs867157175/WRAP73 | 0.00002 | 0.000024 | 0 | 0 | 0.00003 | 0 | 0 | 0.003 |
| rs139430946/ZFHX3 | 0.001 | 0 | 0.00007 | 0 | 0.0005 | 0 | ***0.01*** | ***0.009*** |

**^&^ ,** allele frequencies pointing to low-frequency variants are given in bold and italics font.

**^$ ,^ Regional allele frequencies:** By way of examining the allele frequencies reported in gnomADg V4.1. for middle east and GME_AP, we find 12 of the 27 tightly-replicated MO risk variants as low-frequency variants (MAF≥0.01) and the remaining as rare variants in the region – these are rs78661674/APMAP, rs61746813/ATG2A, rs34584424/*CDK7,* rs148204337/*CFAP43,* rs77052999/**GLB1L3,** rs112145183/KLK14, rs117368247/NOPCHAP1, rs151008831/NUP210, rs57896484/**OR2D2,** rs117898584/PLEKHG1, rs868023411/SLC35B2, and rs139430946/ZFHX3.

**^@ ,^ Global allele frequencies:** Examination of the gnomADg_all V4.1. allele frequencies revealed 9 of the 27 MO variants as low-frequency in global populations. Such variants were rs78661674/*APMAP,* rs61746813/*ATG2A,* rs34584424/*CDK7,* rs113062332/FBF1, rs77052999/**GLB1L3,** rs76348543/KCNMB1, rs117368247/NOPCHAP1, rs57896484/**OR2D2, and** rs61740565/TTC32.

**^%^ ,** Five of the variants exhibited minor allele frequency of >5% in African population and one variant in East Asian population among the gnomADg sample set. These values are indicated by italics and red font. These variants are: African population: rs57896484/OR2D2 (MAF: 11%), rs61746813/ATG2A (MAF: 10.4%), rs76348543/KCNMB1 (MAF: 7.1%), rs77052999/GLB1L3 (MAF: 7%), and rs61740565/TTC32 (MAF: 5.2%); East Asian population: rs78661674/*APMAP.*
